# The impact of spatial connectivity on NPIs effectiveness

**DOI:** 10.1101/2023.10.23.23297403

**Authors:** Chiara E. Sabbatini, Giulia Pullano, Laura Di Domenico, Stefania Rubrichi, Shweta Bansal, Vittoria Colizza

## Abstract

**Background:** France implemented a combination of non-pharmaceutical interventions (NPIs) to manage the COVID-19 pandemic between September 2020 and June 2021. These included a lockdown in the fall 2020 – the second since the start of the pandemic – to counteract the second wave, followed by a long period of nighttime curfew, and by a third lockdown in the spring 2021 against the Alpha wave. Interventions have so far been evaluated in isolation, neglecting the spatial connectivity between regions through mobility that may impact NPI effectiveness.

**Methods:** Focusing on September 2020 – June 2021, we developed a regionally-based epidemic metapopulation model informed by observed mobility fluxes from daily mobile phone data and fitted the model to regional hospital admissions. The model integrated data on vaccination and variants spread. Scenarios were designed to assess the impact of the Alpha variant, characterized by increased transmissibility and risk of hospitalization, of the vaccination campaign and alternative policy decisions.

**Results:** The spatial model better captured the heterogeneity observed in the regional dynamics, compared to models neglecting inter-regional mobility. The third lockdown was similarly effective to the second lockdown after discounting for immunity, Alpha, and seasonality (51% vs 52% median regional reduction in the reproductive number R_0_, respectively). The 6pm nighttime curfew with bars and restaurants closed, implemented in January 2021, substantially reduced COVID-19 transmission. It initially led to 49% median regional reduction of R_0_, decreasing to 43% reduction by March 2021. In absence of vaccination, implemented interventions would have been insufficient against the Alpha wave. Counterfactual scenarios proposing a sequence of lockdowns in a stop-and-go fashion would have reduced hospitalizations and restriction days for low enough thresholds triggering and lifting restrictions.

**Conclusions:** Spatial connectivity induced by mobility impacted the effectiveness of interventions especially in regions with higher mobility rates. Early evening curfew with gastronomy sector closed allowed authorities to delay the third wave. Stop-and-go lockdowns could have substantially lowered both healthcare and societal burdens if implemented early enough, compared to the observed application of lockdown-curfew-lockdown, but likely at the expense of several labor sectors. These findings contribute to characterize the effectiveness of implemented strategies and improve pandemic preparedness.

## BACKGROUND

Non-pharmaceutical interventions (NPIs) represented the primary response to the COVID-19 pandemic in 2020-2021 before mass vaccination campaigns reached a substantial fraction of the population in Europe^1^. After the generalized use of strict lockdowns during the first wave^2–9^, combinations of NPIs reached finer granularity in the second and third waves^10^, occurring in the fall 2020 and in the spring 2021, respectively. These included the closure of certain business sectors (e.g. restaurants, retail and leisure venues), remote education for specific school levels (e.g. high school), bans of gatherings, mobility restrictions, and nighttime curfews at different hours, in addition to less stringent lockdowns. They were meant to manage a rapidly evolving context characterized by the emergence of the first variant of concern^11,12^ and the rollout of vaccination^1^, while pandemic fatigue developed in the population^13–15^.

Spatial heterogeneities in COVID-19 resurgence^16^ and in the geographic seeding of the Alpha variant further added to the complexity of the pandemic phase between the fall 2020 and the summer 2021. In France, the second wave showed a clear spatial pattern with a resurgence in the south-east of the country (**Figure 1a**), likely fueled by summer displacements to touristic destinations. In contrast, the third wave was initially shaped by the seeding of the Alpha variant in the north and in the region of Marseille (Provence-Alpes-Côte d’Azur; **Figure 1a**) then invading other regions through mobility. Population response to nationwide restrictions varied regionally^17,18^ with the potential to affect the epidemiological impact both locally and in other regions connected through mobility fluxes. Spatial connectivity determines geographic spillover events^19–21^ and source-sink mechanisms^22,23^ that can weaken local control policies. Estimating NPIs’ effectiveness and societal burden while accounting for all these elements is key to adequately plan for the medium-term phase of a pandemic, i.e. following the initial emergency and before mass vaccination allows lifting restrictions.

**Figure 1.**
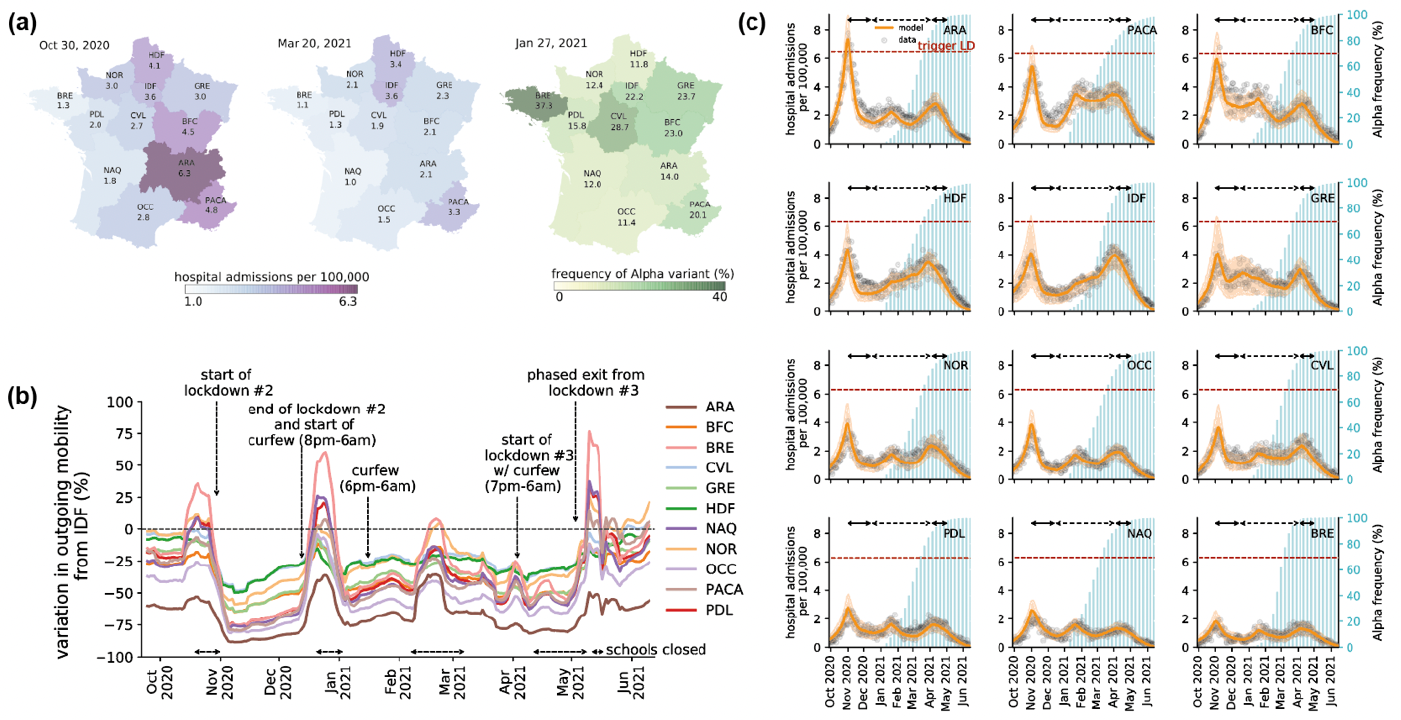
COVID-19 pandemic in French regions between September 2020 and June 2021. **(a)** Regional maps of the per capita hospital admissions as of October 30, 2020 (left, start of the first lockdown) and March 20, 2021 (center, start of the third lockdown in regions IDF, HDF, in one department of NOR and one department of PACA). Hospital admissions displayed on the maps are obtained from a weekly rolling mean of the data. Regional map of the frequency of the Alpha variant (%) as of January 27, 2021 (right, date of the second genomic surveillance survey). Abbreviations refer to the regions: ARA, Auvergne-Rhône-Alpes; BFC, Bourgogne-Franche-Comté; BRE, Brittany; CVL, Centre-Val de Loire; GRE, Grand Est; HDF, Hauts-de-France; IDF, Île-de-France, the region of Paris; NAQ, Nouvelle Aquitaine; NOR, Normandy; OCC, Occitanie; PACA, Provence-Alpes-Côte d’Azur; PDL, Pays de la Loire. **(b)** Variation of regional outgoing mobility from Île-de-France to other regions with respect to pre-pandemic levels. The time intervals indicated over the x-axis refer to (planned or enforced) school closures. **(c)** For each region, the panel shows the model (orange curve and shaded area indicating the median and 95% probability range) fitted to daily hospital admissions data (gray dots). Each plot also shows the percentage of Alpha variant over time (blue histogram, right y-axis). The dashed horizontal line refers to the threshold triggering the second lockdown. Black arrows at the top of each plot correspond to social distancing measures: the second lockdown during the second wave in the fall 2020 (continuous line), followed by the curfew (dashed line) from January to March 2021, and the third lockdown during the third wave in the spring 2021 (continuous line).

Here, we introduced a regionally-based spatially-explicit epidemic metapopulation model that integrates mobility fluxes estimated from mobile phone data to study the COVID-19 pandemic in France between September 2020 and June 2021. Accounting for spatiotemporal heterogeneities, we estimated the effectiveness of implemented NPIs by disentangling spatial and temporal effects (inter-regional mobility, Alpha variant seeding and penetration, seasonality, vaccination). We also examined alternative policy options to the ones implemented by authorities to best balance the epidemiological and healthcare impacts of interventions with the resulting burden of restrictions. The aim was to improve the guidance of policy decisions for the medium-term management of future pandemic threats.

## METHODS

### Restrictions

In **Table 1** we describe the main restrictions implemented in France during the study period (September 2020 – June 2021). In response to the second wave, on October 17, 2020, a nighttime curfew from 9pm to 6am was enforced in several areas with degrading indicators. Due to the rapid surge in the number of infections, a national lockdown was put in place starting October 30, 2020. The restrictions imposed were less stringent compared with the first national lockdown in the spring 2020, as schools and a larger number of job sectors were allowed to remain open. Bars, restaurants, gyms, leisure venues and other non-essential services were closed. Displacements were limited to a maximum radius of one kilometer from home. The lockdown was lifted on December 15, 2020, with the application of a nighttime curfew (8pm – 6am). Soon after the detection of the Alpha variant on the French territory in early 2021, curfew hours were extended nationally between 6pm and 6am on January 16, 2021. Following the rise in cases due to the Alpha epidemic initiating the third wave, on March 20, 2021 localized lockdowns were implemented in the regions of Île-de-France, Haute-de-France and in few other French departments at high incidence. The lockdown was extended to the whole country soon after on April 3, 2021, with the closure of non-essential activities. The gastronomy sector remained closed and the curfew was maintained starting at 7pm. However, differently from the second lockdown, schools were closed for most of the period, extending the planned closure for school holidays of 1 week in the primary schools, and of 2 weeks in the middle and high schools. Movements restrictions were only applied to trips exceeding 10km from the place of residence.

**Table 1.**
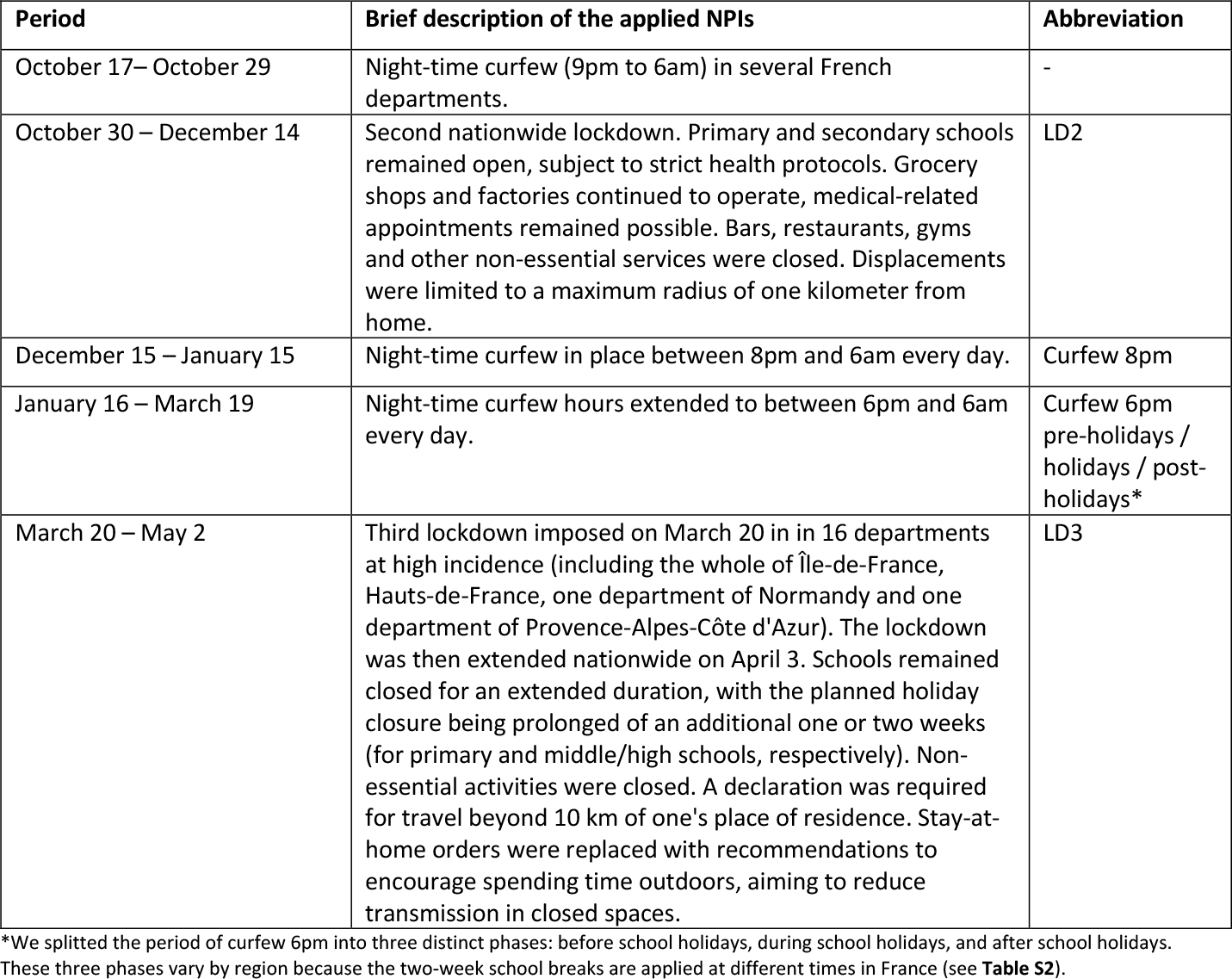
Description of the restrictions applied in France between September 2020 and June 2021.

Also, stay-at-home orders of the second lockdown were converted into recommendations to spend time outdoor to limit transmission in closed settings in this period. The third lockdown ended on May 3, 2021.

### Data

#### Mobility

Anonymized aggregated mobility fluxes extracted from mobile phone signaling data were provided by the Orange business service Flux Vision^18,24^. Data included de-identified origin-destination matrices reporting the daily number of user displacements among 1,436 EPCI (Établissements Publics de Coopération Intercommunale) areas in mainland France. The anonymization procedure was approved by the French data protection authority CNIL (Commission Nationale de l’Informatique et des Libertés). Origin-destination matrices were aggregated at the regional level to compute weekly coupling probabilities *p*_*ji*_ between regions *i* and *j* and inform our model (see the Model subsection). The coupling probability *p*_*ji*_ for a given week is defined as the probability that a resident in *I* visits *j* due to his mobility trajectory:

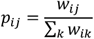

where *w*_*ji*_ is the average number of daily trips between *i* and *j* for a given week. We considered the average to avoid daily fluctuations in the weekly pattern. We chose as a pre-pandemic baseline period the week 6, 2020 (February 03, 2020 – February 09, 2020), as in previous work^17^.

#### Seasonality

A number of studies have suggested that SARS-CoV-2 transmission is seasonally varying, modulated by environmental variables and environmentally-mediated social behavior^25–28^. We integrated seasonality in the regional transmissibility (see Model subsection) based on estimates provided in Ref.^29^. These estimates quantify the impact of seasonal climatic conditions on transmission rate based on daily data from the National Oceanic and Atmospheric Administration (**Figure S12**). We fitted the estimates with a sinusoidal function with 1-year period, one per each region, in order to obtain daily values of the seasonality factor *σ*_*i*_(*t*) affecting transmission in region *i* on day *t*. We used a least-squares optimization function for the fit.

#### Alpha variant

According to genomic surveillance data^30^, the Alpha variant started to circulate in France at the end of 2020 and replaced the previous SARS-CoV-2 strains in March 2021^31^. Results of a large-scale genome sequencing initiative launched in January (so-called Flash surveys^30^) showed that the Alpha variant was responsible for 3.3% of detected COVID-19 cases on January 8, 2021 at the national level, with large spatial heterogeneity, ranging from 0.2% penetration to 6.9%. We modeled the overall virus transmissibility (i.e. wild strain and Alpha variant) by accounting for the regional frequency of Alpha over time (**Figure S12**) and its transmission advantage, to be fitted. In agreement with prior estimates^30^, we found that the SARS-CoV-2 Alpha variant was 58% more transmissible than the wild type in the invasion phase. We also considered a 64% increase in hospitalization rate^32^.

#### Vaccination

We modeled three different vaccination strata, i.e. unvaccinated, vaccinated with one dose or with two doses, based on data on the administration of doses by region^33^ **(Figure S12)**. We assumed vaccines to be effective 14 days after injection. We considered 60% vaccine effectiveness against infection and 15% against transmission after the first injection^34,35^, increasing to 87.5% and 68%, respectively, after the second injection^35,36^. We considered 80% vaccine effectiveness against hospitalization after one dose, and 97,2% after two doses^34,36^. We did not consider waning in vaccine effectiveness in the timeframe under study.

#### Normalcy index

The Economist’s Normalcy index^37^ is a measure of the impact of the pandemic on human behavior, integrating multiple daily indicators of human activities in a score from 0 to 100, with 100 representing the pre-pandemic level (**Figure S1**). We used the Normalcy index to define the effective days under restrictions (see the corresponding subsection).

### Model

#### Metapopulation model summary

We used a discrete stochastic transmission model with a metapopulation structure at the regional level. The population was divided in the 12 regions of mainland France (excluding Corsica). The daily force of infection *λ*_*i*_ in region *i* at time *t* accounts for disease transmission due to (i) infected residents not moving out of the region (*λ*_*ii*_) (ii) infected visitors coming from other regions 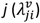 and (iii) returning residents previously infected in other regions 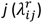^38^:

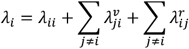

Let *β*_*i*_(*t*) be the transmission rate of region *i* on day *t, p*_*ji*_(*t*) the coupling probability between regions *i* and *j* estimated from mobility data. Let 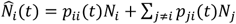 and *Î*_*i*_(*t*) = *p*_*ii*_(*t*)*I*_*i*_(*t*) + ∑_*j*≠*i*_ *p*_*ji*_(*t*)*I*_*j*_(*t*) be the effective population and the effective number of infections in region *i* on day *t*, respectively^38^. Then the daily force of infection can be written as

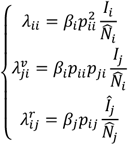

where *β*_*i*_(*t*) accounts for both seasonality and the presence of the Alpha variant. All variables in the previous equations depend on daily time *t*, but we dropped the dependence for the sake of clarity. We indicate with A_*i*_(*t*) ∈ [0,1] the variant’s penetration in region *i* on day *t*, with *η*_*i*_(*t*) the transmission advantage of the Alpha variant, and with *σ*_*i*_(*t*) the seasonality factor. The resulting transmission rate in region *i* on day *t* can be written as:

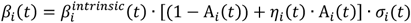

with 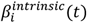 being the fitted transmission rate.

We considered a SEIHR compartmental scheme (**Figure S2** of the **Supporting information, SI**), including susceptible, exposed, infectious, hospitalized and recovered. The model was stratified by three different vaccination status. All the data presented before were integrated in the model at the regional level (**SI**). Parameters, values, and sources used to define the compartmental scheme are listed in **Table S1**. The study period ranges from September 21, 2020 to June 13, 2021 (w39-2020 to w23-2021), to capture the second and third COVID-19 waves.

#### Inference framework and validation

Model parameters were estimated in a Bayesian framework using Markov Chain Monte Carlo (MCMC) method (**SI**). The likelihood function was evaluated on daily data of regional hospital admissions. The log-likelihood function is of the form:

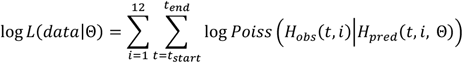

where 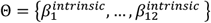 indicates the regional transmission rates to be estimated, *H* (*t, i*) is the observed number of hospital admissions on day *t* in the region *i, H*_*pred*_(*t, i*, Θ) is the number of hospital admissions predicted by the model using parameter values Θ, *Poiss* |⋅ *H*_*pred*_(*t, i*, Θ)O is the probability mass function of a Poisson distribution with mean *H*_*pred*_(*t, i*, Θ), and [*t*_*start*_, *t*_*end*_] is the time window considered for the fit. These time windows are defined based on the interventions applied in France (**Table 1**). When the time window includes a lockdown, we also fitted the time from lockdown implementation to hospitalization peak for each region, to better capture the peak and the decline of the epidemic curve that may vary regionally based on population response^39^ (**Table S3**). We validated the model by comparing its predictions of the percentage of antibody-positive people with seroprevalence estimates from multiple studies at different dates^40,41^. Modeling results were in good agreement with the serological estimates, both at the regional and national levels (**Figures S13, S14, SI subsection 2**.**4**).

### Reproductive numbers

We computed the regional basic reproductive numbers with the next-generation approach^42^ for each time window of the fit. The resulting estimates are obtained from the fitted transmissibility values of the metapopulation model that account for the mobility process. We distinguished between the basic reproductive number R_0_ obtained from the fitted transmissibility *β*_*i*_(*t*) that includes seasonality and the increasing frequency of Alpha, and the intrinsic basic reproductive number R_0_^intrinsic^ that discounts for the seasonal and variant effects, in order to compare different time windows. Analogously, we computed the corresponding effective reproductive numbers, R and R^intrinsic^, accounting for immunity.

### Counterfactual lockdown scenarios

We modeled alternative policy scenarios with respect to the lockdown-curfew-lockdown policy implemented in France, and considered stop-and-go nationwide lockdowns, i.e. repeated lockdowns intercut by periods with no restrictions. As French authorities did not establish thresholds to apply restrictions, we considered stop-and-go lockdown scenarios triggered and released by a given threshold of per-capita hospital admissions, and then we systematically explored these thresholds. We used as reference value *T* of the trigger threshold the hospitalizations per capita in the region at the highest hospitalization incidence when the second lockdown was applied, i.e. the Auvergne-Rhône-Alpes region (ARA). This level corresponds effectively to the highest hospital occupation that authorities deemed sustainable. For the release threshold *R* we considered instead the average hospitalization incidence reported across regions at the moment of lifting the second lockdown. We used the average value because, differently from the triggering threshold, the release threshold is not constrained to a maximum capacity.

We systematically explored different values of trigger and release thresholds, expressed as percentage threshold variations (from *T* to *T*-95%, from *R*+5% to *R*-90%). Given a pair of values of trigger and release thresholds, a nationwide lockdown is activated in the stop-and-go lockdown scenarios when a region first reaches *T*, and it is lifted when the last region reaches *R*. A table with the threshold values is included in the SI (**Table S5**). In the scenarios we simulate a repetition of lockdowns, triggered and lifted according to the above rules, assuming that their stringency would be equal to the one estimated for the second lockdown applied in France (LD2) for the first lockdown in the stop-and-go series, and to the one estimated for the third lockdown applied in France (LD3) for the following lockdowns simulated in the series. This was done to align with the observed political choice of moving from a lockdown largely imposing at-home restrictions (LD2) to one promoting time spent outdoors (LD3). In each simulated nationwide lockdown in the stop-and-go scenarios, the transmissibility and the inter-regional mobility are set to the estimated values of the corresponding lockdown applied in France in the period under study (LD2 or LD3, **Figure S23**). The phasing out of each lockdown was simulated through a two-week piecewise linear function to capture a progressive return to normality after restrictions^43^ (**Figure S22**).

### Effective days under restrictions

We used the Normalcy index^37^ to weigh the days under restrictions by capturing population response and to compare restriction days across intervention scenarios. We defined an “effective day” 𝒟_*t*_ spent under restrictions as

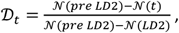

where 𝒩(*t*) is the Normalcy index at time *t*. 𝒟_*t*_ ranges from 0 to 1, with 0 corresponding to the pre-lockdown situation in early October 2020 and 1 representing a day under the second lockdown. We estimated 𝒟_*t*_ =0.58 under curfew (average over all curfew types applied from December 2020 to April 2021) and 𝒟_*t*_=0.77 in the third lockdown. Effective days are also computed for the counterfactual scenarios, based on the duration of implementation emerging from the choice of the trigger and release thresholds.

### Non-spatial model

We tested a non-spatial model, i.e. a model where regions are not coupled by mobility (*p*_*ji*_ = 0). We fitted the model to the regional hospital admission data (**Figure S15**) and evaluated its performance in comparison to the metapopulation model integrating mobility. We performed a model selection test using the deviance information criterion (DIC) and we evaluated the errors of each model using the mean absolute error (MAE) metric (**Subsection 2**.**5 of the SI**).

## RESULTS

In the fall of 2020, French authorities introduced control measures in response to the growing epidemic (**Figure 1, Table 1**). The nighttime curfew implemented in few departments on October 17 was followed by a national lockdown on October 30, the second since the start of the pandemic. Inter-regional mobility dropped by 43-85% in the first three weeks of lockdown compared to pre-pandemic levels, depending on the region (**Figure 1b**). By fitting the metapopulation model to hospital admission data (**Figure 1c**), we estimated a regional median reduction of the basic reproductive number R_0_ of 45% (IQR 42-52%) during the second lockdown compared to the pre-lockdown value in early October 2020 (**Figure 2a**). The 8pm nighttime curfew implemented to phase out the second lockdown in mid-December was not enough to limit community transmission (R>1; **Figure 2b**), due to winter seasonality and Alpha initial spread. Anticipating at 6pm the start of the nighttime curfew on January 16, 2021 resulted in R<1 in all regions except Île-de-France and Hauts-de-France. Such control however deteriorated over time, due to Alpha becoming dominant in the country. Inter-regional mobility remained fairly stable during this time, with a reduction of 23-70% across regions compared to pre-pandemic level, with the exception of the increase registered in February for the school holidays.

**Figure 2.**
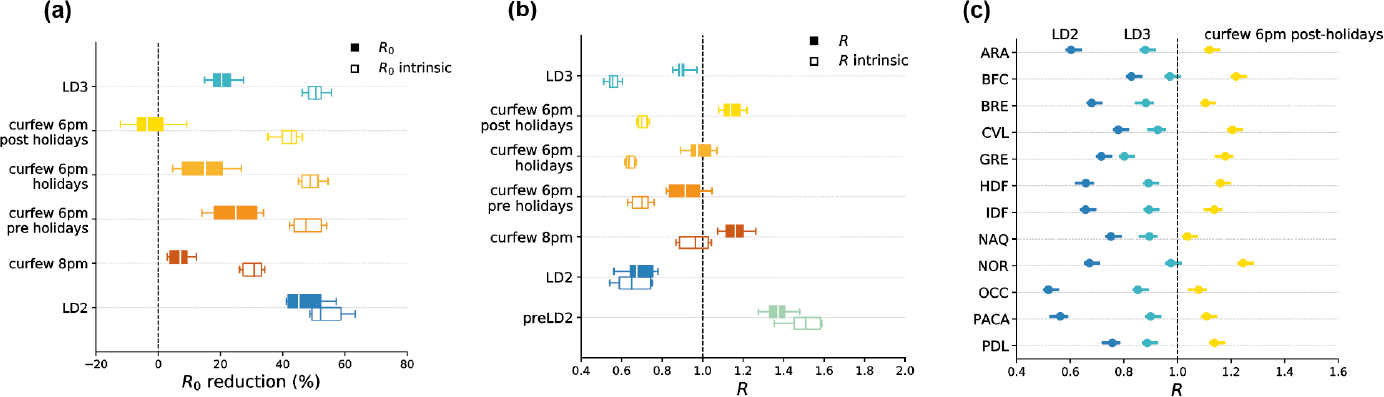
Estimated impact of implemented NPIs. **(a)** Reduction in the estimated regional basic reproductive numbers R_0_ associated to the implemented NPIs compared with the values estimated before the second lockdown. Box plots represent the median (line in the middle of the box), interquartile range (box limits) and 2.5th and 97.5th percentiles (whiskers) of the estimated values for the 12 French regions. Filled boxplots represent reductions estimated by the fit accounting for all time-varying processes (R_0_); void boxplots represent the same reductions discounting the seasonal and Alpha effects (R_0i_^ntrinsic^). **(b)** Estimates of the regional effective reproductive numbers R for the implemented NPIs; box plots as defined in (a). Filled boxplots represent fit estimates accounting for all time-varying processes (R); void boxplots represent the same estimates discounting the seasonal and Alpha effects (R^intrinsic^). **(c)** Regional effective reproductive numbers R for the second lockdown (LD2, dark blue dots), third lockdown (LD3, light blue dots) and the 6pm nighttime curfew in the period following the winter holidays (yellow dots). Dots represent median reproductive number values and error bars the 95% confidence interval. LD2: second lockdown in the fall 2020; curfew 8pm: nighttime curfew starting at 8pm, from mid December 2020 to mid January 2021; curfew 6pm pre-holidays: mid January 2021 to mid February 2021; curfew 6pm holidays: mid February 2021 to late February 2021; curfew 6pm post-holidays: late February 2021 to early April 2021 (see **Table S2**); LD3: third lockdown in the spring 2021.

Discounting for Alpha and seasonality allows us to compare the effectiveness of the 6pm curfew throughout the period in which the variant was becoming dominant, while entering into the spring season. Little change was estimated during the school holidays (R_0_^intrinsic^ reduction of 49% (IQR 46-51%) vs. 48% (IQR 44-52%) in the pre-holiday period), but the effectiveness lowered afterwards (R_0_^intrinsic^ reduction of 43% (IQR 40-44%) post-holiday). Effectiveness varied comparably in all regions during these 3 periods with the 6pm curfew (Spearman correlation r=0.9, p<0.01; **Figure S19**), but less so when comparing 8pm and 6pm curfew periods (r=0.6, p=0.04). With hospital admissions rapidly increasing (estimated regional median R=1.14 (IQR 1.11-1.19)), on March 20 authorities enforced a third lockdown in the highest incidence areas (Île-de-France, Haute-de-France, and few other departments; **Figure 1a**), then extended it nationwide on April 3, till May 3.

The third lockdown resulted in a 6% higher Normalcy index compared to the second lockdown (**Figure S1**). A small mobility drop was registered passing from the curfew to the third lockdown (**Figure 1b**). We estimated a 20% (IQR 18-23%) regional median reduction of R_0_ during the third lockdown (**Figure 2a**), i.e. less than half the value achieved with the application of the second lockdown. By discounting seasonality and the Alpha variant, our model indicates however that the intrinsic effectiveness of the two lockdowns was rather similar (R_0_^intrinsic^ reduction of 52% (IQR 49-59%) in the second lockdown vs. 51% (IQR 48-52%) in the third).

NPI effectiveness varied regionally throughout the period under study (**Figure 2c, Figure 3a**). The metapopulation model integrating observed inter-regional mobility was found to be statistically preferable with respect to a non-spatial model neglecting connectivity (**Table S4**), yielding a lower mean absolute error in 64% of the regions (**Figure S16**). Relative deviations on the estimates of R obtained with a non-spatial model compared to the metapopulation approach could be >40% and were found to increase with increasing mobility for a given regional population (Spearman correlation r=0.46, p< 10^−4^, **Figure 3b**).

**Figure 3.**
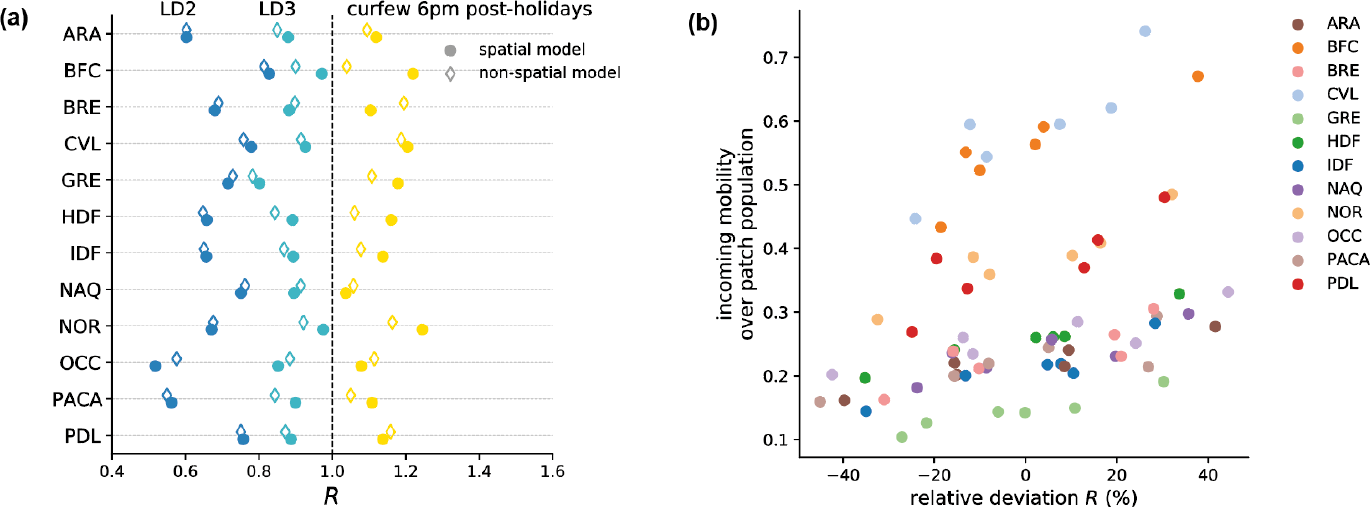
Spatial vs. non spatial model. **(a)** Regional effective reproductive numbers R for the second lockdown (LD2, dark blue dots), third lockdown (LD3, light blue dots) and the 6pm nighttime curfew in the period following the winter holidays (yellow dots). Dots represent median reproductive number values. Filled symbols refer to the estimates obtained with the spatial model, void symbols represent the estimates obtained with the non-spatial model, i.e. neglecting inter-regional mobility. **(b)** Scattered plot between the incoming mobility divided by the patch population and the relative deviation in the estimated effective reproductive numbers obtained with the non-spatial model, for the different NPIs applied (six dots for each region, referring to six different NPIs). Colors of the dots refer to the regions. Results of a Spearman correlation test (r=0.46, p-value < 10^−4^). LD2: second lockdown in the fall 2020; curfew 8pm: nighttime curfew starting at 8pm, from mid December 2020 to mid January 2021; curfew 6pm pre-holidays: mid January 2021 to mid February 2021; curfew 6pm holidays: mid February 2021 to late February 2021; curfew 6pm post-holidays: late February 2021 to early April 2021 (see **Table S2**); LD3: third lockdown in the spring 2021.

Alpha spread was estimated to be responsible for 129,335 (IQR 112,290-144,396) hospitalizations in mainland France, corresponding to 41% of the overall hospitalizations recorded in the study period (**Figure S10**). Our model predicted that Île-de-France, the region of Paris, was the most impacted by the variant (50% of the overall hospitalizations), followed by Hauts-de-France (48%). The least impacted was Nouvelle Aquitaine (29%). This result was not exclusively explained by the geographical seeding of Alpha (**Figure 1a**). Indeed, if Île-de-France reported the largest variant frequency at the start of January 2021 (**Figure S20**), such ranking was rapidly altered by mid-January, despite the same control measures were being applied nationwide. Vaccination was estimated to prevent 255,195 (IQR 224,993-279,502) hospitalizations in mainland France in the time period under study, equal to 81% of the hospitalizations that were actually reported (**Figure S20**). An anticipated and faster vaccination rollout, as implemented in the UK, would have prevented additional 122,877 (IQR 107,404-137,971) hospitalizations (i.e. additional 39%). Most importantly, without vaccination, implemented NPIs would have not been sufficient to control the Alpha variant (**Figure S20c**).

The application of nighttime curfew allowed authorities to manage the pandemic between the second and third waves, albeit maintaining a high incidence of cases and hospitalizations. To examine whether additional policies could have been more beneficial, we explored counterfactual scenarios with stop-and-go lockdowns. With the trigger and release thresholds (*T* and *R* in **Figure 4** and **Figure 5**; see also **Methods**) computed from the experience of the second lockdown in France in the fall 2020, three lockdowns would have been needed to manage the pandemic between September 2020 and June 2021, reducing by 22% the effective days under restrictions, but increasing hospitalizations by 40%. Also, the impact on regional healthcare would have largely varied, with Bretagne, for example, predicted to have an increase of 190% of its hospitalizations (**Figure 5c**). The second of the three lockdowns foreseen under this scenario would have lasted more than 3 months to control the rise of the Alpha wave (January– April 2021; **Figure 5b**).

**Figure 4.**
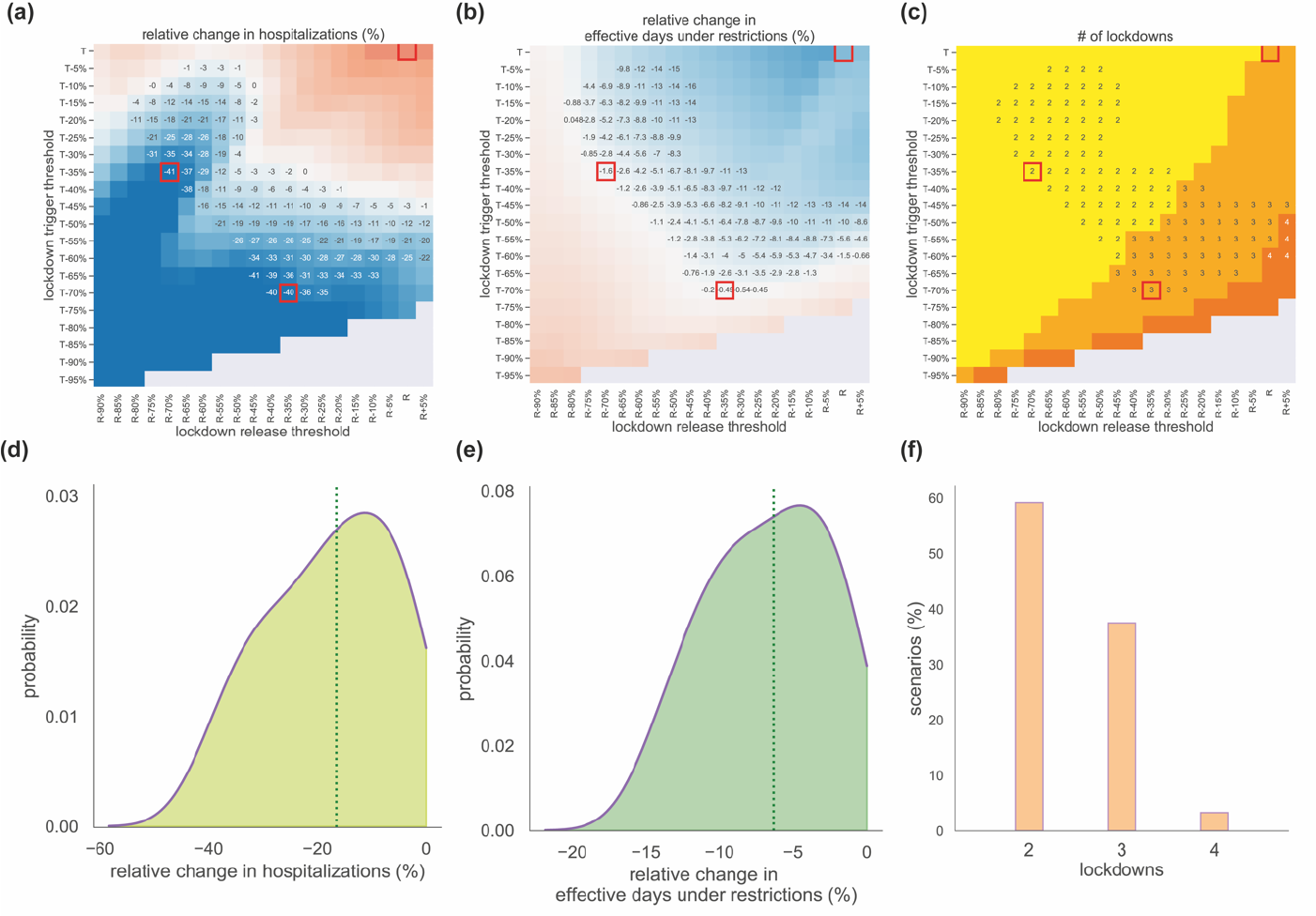
Impact of stop-and-go lockdown scenarios on hospitalizations, effective days under restrictions, and number of lockdowns. **(a-c)** Heatmaps showing the relative variation in cumulative hospital admissions **(a)**, the relative variation in effective days under restrictions **(b)**, and the number of lockdowns **(c)**, as functions of the thresholds for trigger (y-axis) and release (x-axis) of nationwide lockdowns. Relative changes are computed with respect to observations. The red squares indicate specific values of trigger and release threshold that are discussed in the main text and presented in detail in Figure 5. Numerical values are reported only in the area where both hospitalizations and effective days under restrictions are reduced by the lockdowns compared to observations. **(d,e)** Probability distributions of the relative variations in hospitalizations **(d)** and in effective days spent under restrictions **(e)** compared to observations, in the region of the trigger-release parameter space where both quantities are reduced by the lockdowns. The vertical dashed lines represent the median values of the distribution. **(f)** Histogram of the percentage of scenarios with a given number of lockdowns, among the scenarios that reduce both hospitalizations and effective days.

**Figure 5.**
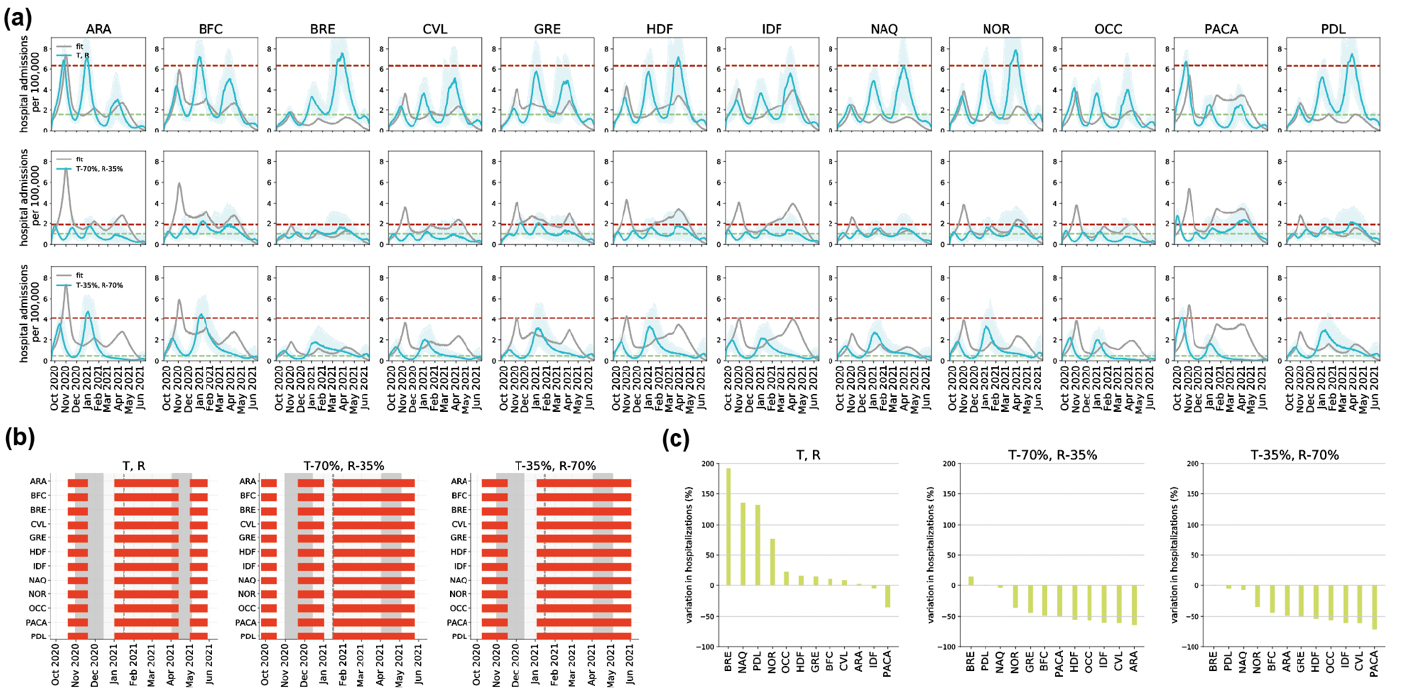
Regional trajectories of stop-and-go lockdown scenarios for specific trigger and release thresholds. **(a)** Trajectory of regional daily hospital admissions, model fit vs. stop-and-go lockdown scenarios for three different choices of the trigger and release thresholds, indicated by the dashed horizontal lines (top: T,R; center: T-70%,R-35%; bottom: T-35%,R-70%). **(b)** Regional timeline of lockdowns, observed (gray areas) vs. lockdown scenarios (red bars). Gray shaded areas in the plots correspond to social distancing measures: the second lockdown during the second wave in the fall 2020 (darker gray), followed by the curfew (lighter gray) from January to March 2021, and the third lockdown during the third wave in the spring 2021 (darker gray). The vertical dashed line denotes the anticipation of the nighttime curfew at 6pm, on January 16, 2021. **(c)** Variation in hospitalizations by region under the lockdown scenario compared to observations

Reducing the trigger and release thresholds, i.e. applying and lifting the lockdowns at lower per capita hospital admissions than observed, would have decreased hospitalizations (**Figure 4a, Figure 5a**) and increased the effective days under restrictions (**Figure 4b**), through two to four lockdowns (**Figure 4c**). For example, maintaining the same number of effective days under restriction as observed, our model predicts that it could have been possible to largely reduce national hospitalizations by around 40% through the early application of two (e.g. with *T-35%* and *R-70%* thresholds) or three lockdowns (e.g. with *T-70%* and *R-35%* thresholds). However, this would have been achieved with a long uninterrupted lockdown period (>4 months; **Figure 5b**).

The interface between decreasing hospitalizations and increasing restriction days yields an intermediate region of threshold values where both quantities are reduced compared to observations, thus limiting both the healthcare and societal burdens. Adopting these criteria, a higher benefit would be on average achieved in the reduction of hospital patients (17%, IQR 9-27%; **Figure 4d**) compared to the reduction of effective days under restrictions (6%, IQR 3-10%; **Figure 4e**), and with a benefit for more than 70% of the regions (**Figure S21**). This would be obtained with two lockdowns in most of the cases (59% of the scenarios), whereas higher lifting thresholds would induce three (38% of scenarios) or four (3%) lockdowns (**Figure 4f**).

## DISCUSSION

Using an epidemic metapopulation model integrating time-varying inter-regional mobility and spatial effects, we provided a detailed analysis of the impact of different measures applied in France between September 2020 to June 2021. Despite their different nature, we showed that the third lockdown (spring 2021) was similarly effective to the second lockdown (fall 2020), after discounting for the transmissibility of the circulating variants, immunity, and seasonal effects. We found a strong difference in the estimated impact of the nighttime curfew starting at 8pm or 6pm, with the latter being able to considerably reduce community transmission (coupled with gastronomy and leisure sectors closed). Under the observed vaccination campaign and NPIs, Alpha was estimated to be responsible for 41% of observed hospitalizations. Conversely, without vaccines, we found that implemented measures would not have been enough to control the Alpha wave. Finally, stop-and-go lockdowns triggered early enough would have resulted in lower hospitalizations and effective days under restrictions compared to the observed lockdown-curfew-lockdown.

Our analysis demonstrates that the spatial model better captures the observed regional dynamics compared to non-spatial models that neglect case importations across regions coupling regional epidemics (**Figure S15, Table S4**). This finding highlights the importance of considering mobility and spatial data to better characterize epidemic transmission processes and evaluate interventions. Even if these interventions are applied nationwide, local conditions (e.g. incidence, penetration of a variant of concern, seasonal effects, human response to interventions) can be rather heterogeneous geographically, so that spillover events across different areas have unexpected implications for the local epidemic dynamics and control^19,20^. We found that reproductive number estimates were largely affected by the spatial connectivity fueling regional epidemics, with relative deviations that on average may be larger than 40% and generally higher for higher shares of incoming mobility compared to the number of inhabitants (**Figure 3b**). Careful considerations should therefore be given to local estimates neglecting the impact of continuous importations from outside areas.

Our analysis confirms the large effectiveness of lockdowns in controlling transmission, in line with prior works^5,7,8,44,45^. Despite their difference in the granularity and definition of restrictions (stay-at-home orders with schools open during the second lockdown vs. stay-outdoor recommendations with extended school holidays in the third lockdown), the two lockdowns implemented in France in the period under study had a similar impact on the epidemic. They reduced the intrinsic transmissibility by 52% and 51%, respectively (**Figure 2a**), compared to the situation in October 2020 for a rather similar Normalcy index. Our findings therefore suggest that the higher effective reproductive number reached during the third lockdown in France compared to the second (R=0.89 vs. R=0.67, respectively; **Figure 2b**) was mainly the result of the Alpha variant spread, characterized by a higher transmissibility, and not of different stringency of restrictions or lower adherence of the population^31^. Similar transmission reductions were estimated for the most stringent tier applied in Italy in the same period (52% reduction)^46^, and corresponding to Normalcy index values close to the ones of the French lockdowns.

We produced new evidence on the effectiveness of intermediate-stringency NPIs, such as nighttime curfews, for which there was little available literature^31,47,48^. Coupled with the closure of the gastronomy sector, the curfew starting at 6pm was found to be considerably effective, suggesting that a moderate intervention focusing restrictions on certain sectors and times of the day may be a viable control option while ensuring a larger functioning of the economy. Over a longer period of time, however, we found that curfews lost effectiveness in all regions (**Figure 2b**), suggesting that pandemic fatigue^13–15^ likely settled in the population. Maintaining it for a long time (in France it was implemented for a total of 188 consecutive days) should therefore be evaluated in light of expected population adherence^15^ and the potential increase in the prevalence of mental health issues^15^. Regional responses changed in a similar way to changing NPIs, but to a lesser degree when comparing the nighttime curfew starting at 8pm with the one starting at 6pm. This suggests that anticipating the start of the curfew required different organizations of daily routines which may be specific to the regional contexts. Prior work already pointed out the role of local socio-economic factors and labor structure in driving the response to restrictions^17,18,49^. This also limits the generalizability of our curfew results to other societal contexts, as it will depend on local social habits involving mixing activities that may be efficiently restricted by the curfew.

The first half of 2021 witnessed a race between the rollout of vaccines and the spread of the Alpha variant. We found that both vaccines and NPIs were key to control the Alpha wave^50^. Specifically, without vaccines, stricter measures should have been adopted to avoid hospital saturation (**Figure S18**).

Other countries opted for different policies, repeating lockdowns. We showed that stop-and-go lockdowns (intercut with periods of no restrictions) could have achieved a substantial reduction of hospitalizations (−40%) for similar number of effective days under restrictions compared to the policy implemented in France, i.e. the application of two lockdowns intercut with a long period of curfew (**Figure 4, Figure 5**). This result would require however acting early, at low hospitalization incidence^43,51–53^. In the balance between ensuring epidemic control and limiting societal impact, we also found a range of thresholds to trigger and release lockdowns that would reduce both hospitalizations and overall effective days under restrictions. However, in the pandemic phase characterized by a more transmissible and severe variant, this would translate in the implementation of a rather long second lockdown (to compensate for the absence of the curfew in between lockdowns), raising again issues of sustainability and acceptance^14,15^ (**Figure 5**).

Our work has a number of limitations due to simplifying assumptions in our analysis. First, we did not consider the age structure of the population, asymptomatic transmission, or changes of travel behavior when infectious, similarly to what commonly done in COVID-19 metapopulation models^43,54^ where the complexity of the model lies in its spatial dimension. Age-specific mixing and the impact of asymptomatic transmission and travel avoidance behavior are effectively absorbed in the estimate of the regional transmissibility and may be a factor behind resulting regional variations^17,55^. Second, in the stop-and-go lockdown scenarios, we considered a 2-week relaxation to phase out restrictions^43^, reproducing what happened in France. Other countries opted instead for more structured tiered systems to guarantee a better control in lifting interventions^46,56^. Also, the thresholds considered to trigger and lift interventions in the scenarios are based on per-capita hospital admissions, implicitly assuming equal hospital capacity across regions. While regional variations exist, the crisis also showed a certain flexibility in adjusting such capacity according to needs^7^. Finally, we used the Normalcy index to define the effective days under restrictions and compare interventions of different stringencies. Other indicators can be defined using the mobility data, which was at the core of restrictions, as we did in prior work^15^. Different indicators should instead be used for a more comprehensive analysis that may include also economic aspects and the impact on mental health whose prevalence was found to increase substantially throughout the curfew in France^15^.

## CONCLUSIONS

Our analysis provides a detailed overview of the epidemiological impact of the various NPIs and of the vaccination campaign implemented in France from September 2020 to June 2021. Using a spatially-explicit regional metapopulation model allows us to disentangle the effects of spatial and temporal drivers – seasonality, Alpha variant geographic seeding and penetration over time, vaccination rollout, time-varying inter-regional mobility – in the estimates of the effectiveness of lockdowns and curfews of different type. Our findings help the design of preparedness plans for the medium-term management of respiratory virus pandemics.

## Supporting information

Supplementary Infromation

## Data Availability

Mobile phone data are proprietary and confidential. We obtained access to these data from the Orange Business Service Flux Vision within the framework of the research project ANR EVALCOVID-19 (ANR-20-COVI-0007). Access to the mobility data can be requested from Orange on a contractual basis. Hospitalization data were obtained from the SIVIC dataset57. Estimates for seasonal transmissibility were obtained from the authors of Ref.29. Vaccination data are available from the French Government data hub (https://www.data.gouv.fr/en/datasets/donnees-relatives-aux-personnes-vaccinees-contre-la-covid-19-1/); the normalcy index is available from The Economist (https://www.economist.com/graphic-detail/tracking-thereturn-to-normalcy-after-covid-19); and Alpha variant penetration data are published in the scientific literature (https://pubmed.ncbi.nlm.nih.gov/33663644/).

## ACKNOWLEDGMENTS

We thank Mélanie Prague for sharing the estimates on seasonal transmissibility. We thank Pierre-Yves Boëlle for useful discussions.

## AUTHORS’ CONTRIBUTORS

VC conceived and designed the study. CES and GP developed the code. CES ran the simulations and analyzed the data. All authors interpreted the results. CES and VC wrote the initial manuscript draft. All authors edited and approved the final version of the Article.

## AVAILABILITY OF DATA AND MATERIALS

Mobile phone data are proprietary and confidential. We obtained access to these data from the Orange Business Service Flux Vision within the framework of the research project ANR EVALCOVID-19 (ANR-20-COVI-0007). Access to the mobility data can be requested from Orange on a contractual basis. Hospitalization data were obtained from the SIVIC dataset^57^. Estimates for seasonal transmissibility were obtained from the authors of Ref.^29^. Vaccination data are available from the French Government data hub (https://www.data.gouv.fr/en/datasets/donnees-relatives-aux-personnes-vaccinees-contre-la-covid-19-1/); the normalcy index is available from The Economist (https://www.economist.com/graphic-detail/tracking-thereturn-to-normalcy-after-covid-19); and Alpha variant penetration data are published in the scientific literature (https://pubmed.ncbi.nlm.nih.gov/33663644/).

## ETHICS APPROVAL AND CONSENT TO PARTICIPATE

Not applicable.

## CONSENT FOR PUBLICATION

Not applicable.

## COMPETITING INTERESTS

We declare no competing interests.

