## Supplementary Infromation for "The impact of spatial connectivity on NPIs effectiveness"

### 1. Normalcy and Stringency index

In **Figure S1** we show two indicators, the Normalcy and the Stringency index, for France and four other European countries. The Economist's Normalcy index[1] and the Stringency index[2] are two measures used to evaluate the impact of the pandemic on human behavior and government policies.

The Normalcy index tracks eight different variables (sports attendance, time at home, traffic congestion, retail footfall, office occupancy, flights, film box office and public transport) to quantify an overall score. The pre-pandemic activity level was set at a Normalcy index of 100 to ease comparison. In the period including the second lockdown, the curfew, and the third lockdown, the index for France was computed to be between 37 and 64. The Stringency index quantifies the intensity of government policies. **Figure S1** shows that in the period under study Normalcy and Stringency index took complementary values, suggesting a duality between the two indicators. In the main analysis we use the Normalcy index instead of the Stringency index because it captures not only the stringency of interventions but also the behavioral response.

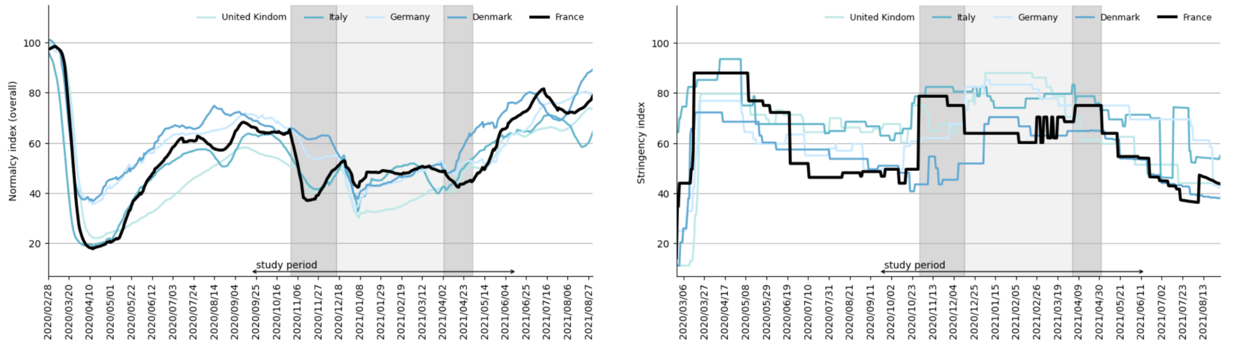

**Figure S1. Indicators. (left)** Normalcy index over time for UK, Italy, Germany, Denmark and France. Shaded rectangle in the plot corresponds to the period of the study. **(right)** Stringency index over time for UK, Italy, Germany, Denmark and France. Shaded areas in the plot corresponds to the period of the second lockdown, third lockdown (dark grey) and night-time curfew in between (light grey). The horizontal arrow highlights the study period.

### 2. SARS-CoV-2 transmission model

#### 2.1. Compartmental model and parameters

**Figure S2** shows the compartmental scheme used to describe COVID-19 disease progression. Individuals are divided into susceptible (S), exposed (E), infectious (I), hospitalized (H) and recovered (R). Hospitalized patients are quarantined and they do not transmit the infection. Parameter values related to infection due to historical strains are reported in **Table S1**. We used estimates available in the literature to inform the average durations of the latency, infectious, and hospitalized compartments in the current work. More in detail:

We set the average latency period to $\epsilon^{-1}$=3.7 days, based on the estimated average length of the incubation period (5.2 days, from Ref.[3]) and discounting the estimated average period between onset of infectiousness and onset of symptoms (1.5 days, from Ref.[4] , computed from the estimates of Ref.[5]). We set the average infectious period to $\mu^{-1}$=2.9 days to match the estimate of the generation time (6.6 days, from Ref.[6]). We set the average time spent in the hospital to 21 days from Ref.[4], and a time lag of 3.6 days was introduced to delay the entry into the hospitalization compartment to align with the estimates on time from onset to hospital admission[7].

The complexity of the metapopulation approach with time-varying mobility coupling the French regions required some simplifications in the compartmental structure. For example, we did not consider asymptomatic or presymptomatic transmission. Such simplifications were commonly adopted in other metapopulation models used to study the COVID-19 pandemic[8,9].

Parameters values for the Alpha variant are presented in the **Methods** section of the main text.

When vaccination starts, we assumed doses to be distributed to either susceptible or recovered individuals with equal probability. Vaccination effectiveness is described in the **Methods** section and reported in **Table S1**.

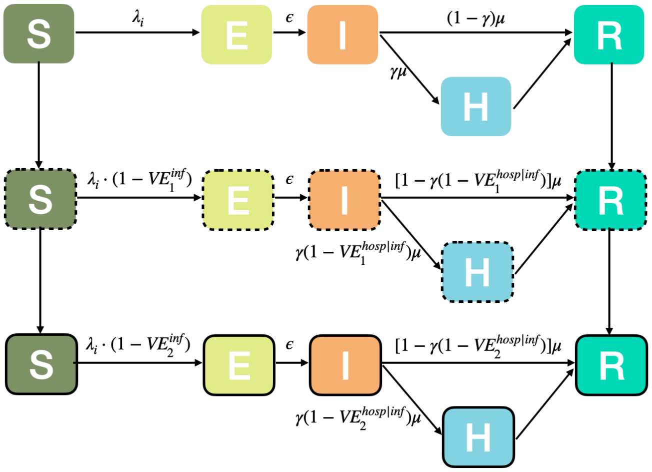

**Figure S2. Compartmental scheme with vaccination.** Compartments with no-lines (top) account for infections due to historical strains for non-vaccinated people. Analogous compartments are considered for vaccinated individuals (dashed and solid lines). S=Susceptible, E=Exposed, I= Infectious, H=severe case admitted to the hospital, R=Recovered. $VE^{hosp|inf}$ has been computed using the relationship: $1 - VE^{hosp} = (1 - VE^{inf}) \cdot (1 - VE^{hosp|inf})$. $VE^{transm}$ is embedded in the force of infection. Variables are defined in **Table S1**

**Table S1.** Parameters, values, and sources used to define the compartmental model for infection due to historical strains.

| Name | Description | Value | Source |
|---|---|---|---|
| $\epsilon^{-1}$ | Latency period | 3.7 days | 3 |
| $\mu^{-1}$ | Infectious period | 2.9 days | 6 |
| $\gamma$ | If infected, probability of going to hospital | 2.1% | 10 |
| $VE_1^{inf}, VE_2^{inf}$ | Vaccine effectiveness in preventing infection in case of 1 or 2 doses | 60%, 87.5% | 11,12 |
| $VE_1^{hosp}, VE_2^{hosp}$ | Vaccine effectiveness in preventing hospitalization in case of 1 or 2 doses | 80%, 97.2% | 11,12 |
| $VE_1^{transm}, VE_2^{transm}$ | Vaccine effectiveness in preventing transmission in case of 1 or 2 doses | 15%, 68% | 13 |

#### 2.2. Inference framework

The time windows used for the fit are defined based on the interventions applied in France (**Table 1, Table S2**), specifically: pre-second lockdown (Sept 21,2020 – region dependent date), second lockdown (region dependent date – Nov 22, 2020), curfew 8pm (Dec 15, 2020 – Jan 15, 2021), curfew 6pm pre holidays (Jan 16, 2021 – region dependent date due to school holidays), curfew 6pm during holidays (region dependent date due to school holidays), curfew 6pm post holidays (region dependent date due to school holidays – April 3, with the exception of few departments), third lockdown (April 3, with the exception of few departments – Jun 13, 2021).

**Table S2.** Time windows corresponding to curfew and holidays.

| NPIs | Time window used | Regions |
|---|---|---|
| Curfew 6pm pre holidays | Jan 16, 2021 – Feb 6, 2021 | ARA,BFC,NAQ |
| | Jan 16, 2021 – Feb 20, 2021 | BRE,CVL,GRE,HDF, NOR,PACA, PDL |
| | Jan 16, 2021 – Feb 13, 2021 | IDF,OCC |
| Curfew 6pm holidays | Feb 7, 2021 – Feb 21, 2021 | ARA,BFC,NAQ |
| | Feb 21, 2021 – Mar 7, 2021 | BRE,CVL,GRE,HDF, NOR,PACA, PDL |
| | Feb 14, 2021 – Feb 28, 2021 | IDF,OCC |
| Curfew 6pm post holidays | Feb 22, 2021 – Apr 3, 2021 | ARA,BFC,NAQ |
| | Mar 8, 2021 – Apr 3, 2021* | BRE,CVL,GRE,HDF, NOR,PACA, PDL |
| | Feb 29, 2021 – Apr 3 2021* | IDF,OCC |

*third lockdown applied earlier (on Mar 20, 2021) in IDF, HDF, PACA.

The model was initialized on March 1, 2020 from estimated prevalence obtained from prior modeling work[14], which was validated against serological data. Each stochastic run of the metapopulation model was initialized on that date by sampling the prevalence of each compartment from a Gumbel-r distribution. Its cumulative distribution function is given by:

$$F(x;\mu,\beta) = e^{-e^{-\frac{(x-\mu)}{\beta}}}$$

where β is the scale parameter, which controls the spread or dispersion of the distribution; μ is the location parameter, which represents the location of the distribution on the x-axis. In our analysis, we opted for the Gumbel distribution as it resulted to be the best distribution in terms of AIC among a set of commonly used distributions (Alpha, Gamma, Invgamma Levy, Loggamma, Lognorm, Powernorm, Norm) when fitting the prevalence estimated in our prior work[14] through a maximum likelihood approach. We chose this initialization procedure as surveillance data prior to March 1, 2020 were of lower quality given that the surveillance system for hospitalizations was being developed, and this hindered the fitting procedure of the metapopulation model prior to the exponential increase of cases. In addition, fitting the early start of the pandemic requires also fitting the seeding date in each region, i.e. potentially in the month of December 2019. However, we lack mobility data for the end of 2019, therefore we were forced to use a region-specific model for the fit of the early phase, without spatial connectivity, and then use its results to initialize our metapopulation model. We expect this not to impact the successive spatial dynamics because of the small epidemic size prior to March 1, 2020 and of the long time period between the initialization (March 2020) and the start of the period under study (September 2020). This is further supported by the validation of the model at subsequent dates and by the model selection analysis illustrating how the metapopulation model better describes the observed dynamics compared to a non-spatial model.

The metapopulation model fitted the epidemic trajectories of hospital admission data from March 1, 2020 to June 13, 2021.

Model parameters were estimated in a Bayesian framework by sampling the posterior parameter distribution obtained by updating prior beliefs based on a likelihood function. The likelihood function is evaluated on daily data of regional hospital admissions **(Methods** section *Inference framework and validation***)**. We used Markov Chain Monte Carlo (MCMC) to obtain posterior distributions, assuming a uniform prior. We used three independent chains, with each chain performing 3000 steps, to approximate the posterior distribution. We used the Metropolis-Hasting algorithm to accept or reject the set of parameters at each step. We performed 200 stochastic simulations to compute median values and associated 95% probability ranges for all quantities of interest.

To demonstrate that our model is able to estimate the parameters with the proposed inference approach and does not suffer from identifiability issues, we performed the following synthetic experiment. We parameterized the model using as priors the set of parameter values estimated with the first MCMC, and we re-calibrated the model with another MCMC procedure. By retrieving the same set of parameter values, we showed that the model was well identified and could be calibrated without bias.

#### 2.2.1. Parameters fitted in the MCMC procedure

We used daily hospital admission data at the regional level to fit the model.

The values of the fitted transmission rates ($\beta_i^{intrinsic}(t)$) and time from lockdown implementation to hospitalization peak are reported in **Table S3.** The distributions of the fitted parameters are reported in **Figures S3-S11**

**Table S3. Values of the fitted parameters.** The parameter β refers to the intrinsic transmission rate $\beta_i^{intrinsic}$ defined in the main text.

| Region | β(pre-LD2) | β(LD2) | β(curfew 8pm) | β(curfew 6pm pre holidays) | β(curfew 6pm holidays) | β(curfew 6pm post holidays) | β(LD3) | Time from lockdown application to hospitalization peak, LD2 | Time from lockdown application to hospitalization peak, LD3 |
|---|---|---|---|---|---|---|---|---|---|
| **ARA** | 0.573627 | 0.218227 | 0.377573 | 0.291390 | 0.292125 | 0.325563 | 0.290536 | 1 days | 8 days |
| **BFC** | 0.625011 | 0.315116 | 0.459515 | 0.338697 | 0.328621 | 0.386210 | 0.340951 | 2 days | 5 days |
| **BRE** | 0.481102 | 0.242640 | 0.328915 | 0.221431 | 0.234657 | 0.272123 | 0.239864 | 6 days | 0 days |
| **CVL** | 0.539905 | 0.280794 | 0.398681 | 0.298373 | 0.288718 | 0.311664 | 0.281231 | 4 days | 5 days |
| **GRE** | 0.576878 | 0.27459 | 0.364502 | 0.333694 | 0.316897 | 0.374449 | 0.280219 | 2 days | 0 days |
| **HDF** | 0.575018 | 0.250048 | 0.385831 | 0.327443 | 0.295461 | 0.323445 | 0.274093 | 2 days | 6 days |
| **IDF** | 0.548415 | 0.260598 | 0.394023 | 0.338249 | 0.337227 | 0.355278 | 0.320925 | 0 days | 12 days |
| **NAQ** | 0.532347 | 0.273099 | 0.385755 | 0.261476 | 0.263212 | 0.285782 | 0.267691 | 3 days | 1 days |
| **NOR** | 0.572577 | 0.241925 | 0.377648 | 0.259662 | 0.259865 | 0.307323 | 0.264029 | 2 days | 9 days |
| **OCC** | 0.567135 | 0.190596 | 0.393210 | 0.272494 | 0.258899 | 0.274563 | 0.254088 | 1 days | 8 days |
| **PACA** | 0.598687 | 0.217141 | 0.465066 | 0.332263 | 0.324089 | 0.340098 | 0.324295 | 2 days | 8 days |
| **PDL** | 0.542186 | 0.276619 | 0.374060 | 0.248372 | 0.262255 | 0.308712 | 0.269117 | 3 days | 0 days |

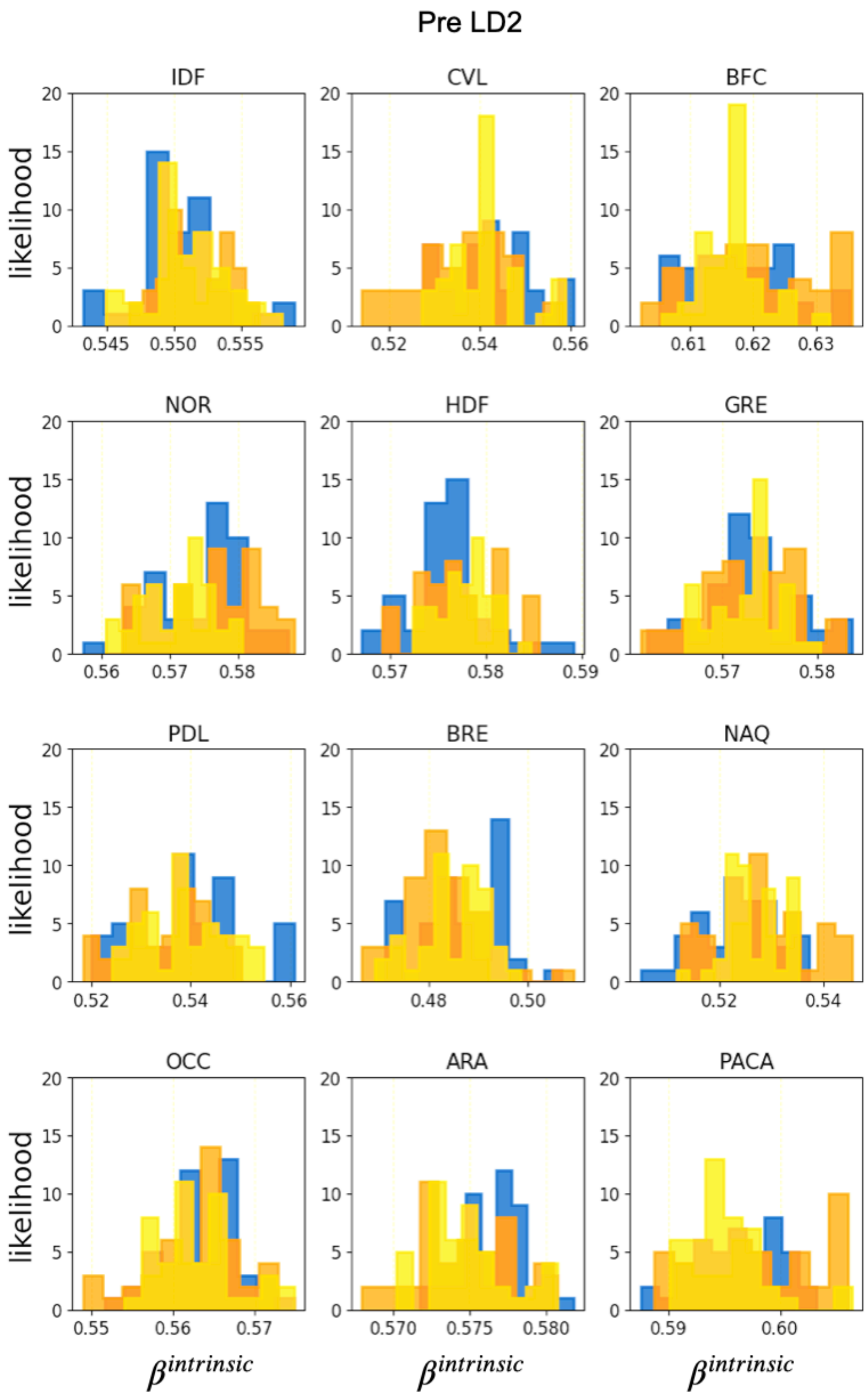

**Figure S3. Histograms of samples.** We reported the histograms of sampled values for the $\beta^{intrinsic}(pre\ LD2)$ parameter in each region, obtained from three independent chains (yellow, orange and blue histogram), after discarding the burn-in period.

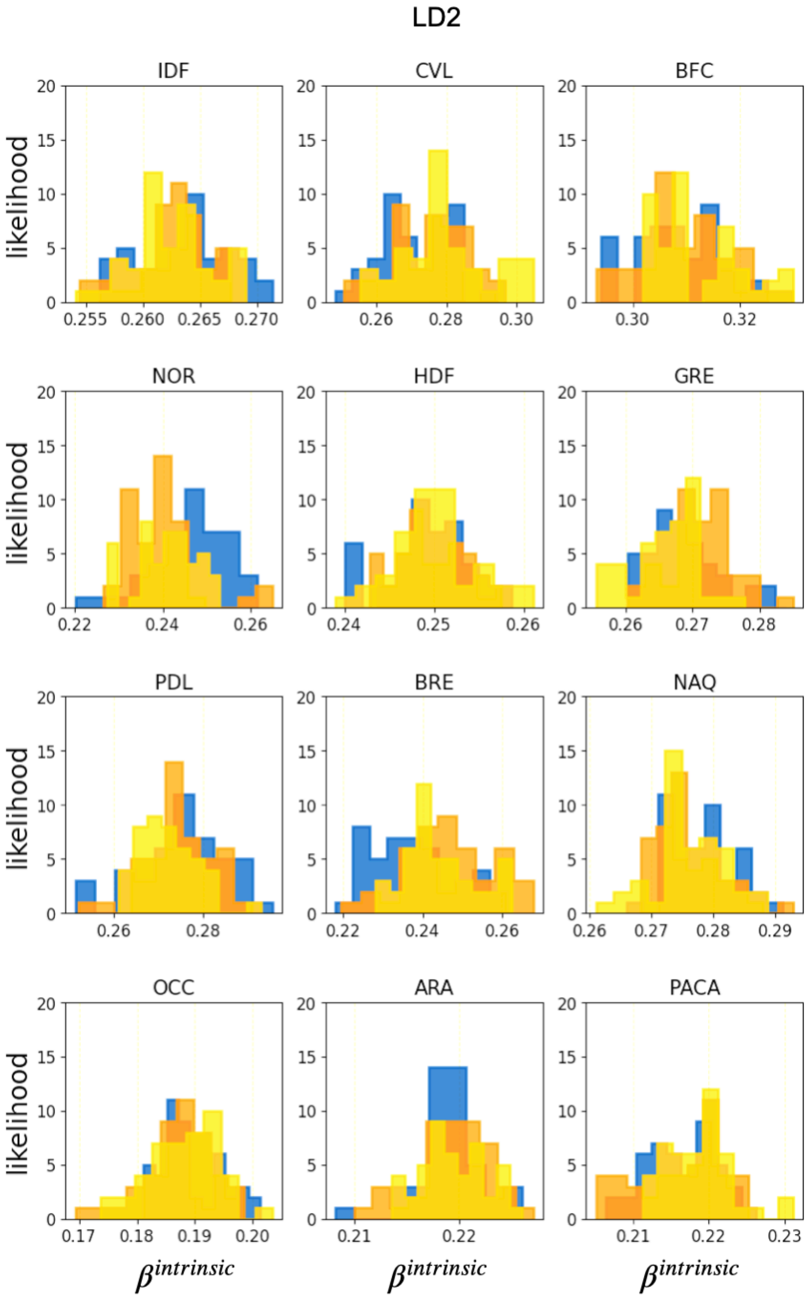

**Figure S4. Histograms of samples.** We reported the histograms of sampled values for the $\beta^{intrinsic}(LD2)$ parameter in each region, obtained from three independent chains (yellow, orange and blue histogram), after discarding the burn-in period.

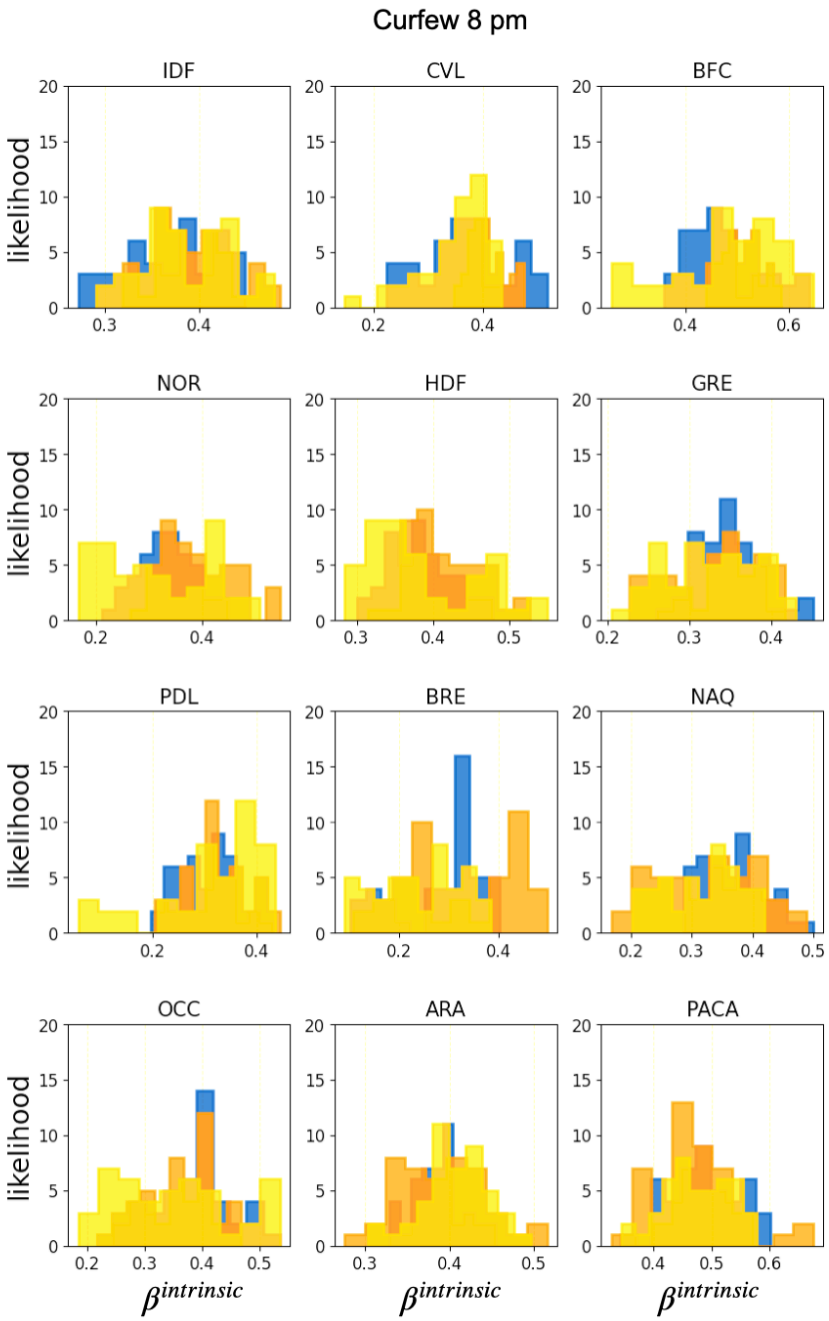

**Figure S5. Histograms of samples.** We reported the histograms of sampled values for the $\beta^{intrinsic}(curfew\ 8pm)$ parameter in each region, obtained from three independent chains (yellow, orange and blue histogram), after discarding the burn-in period.

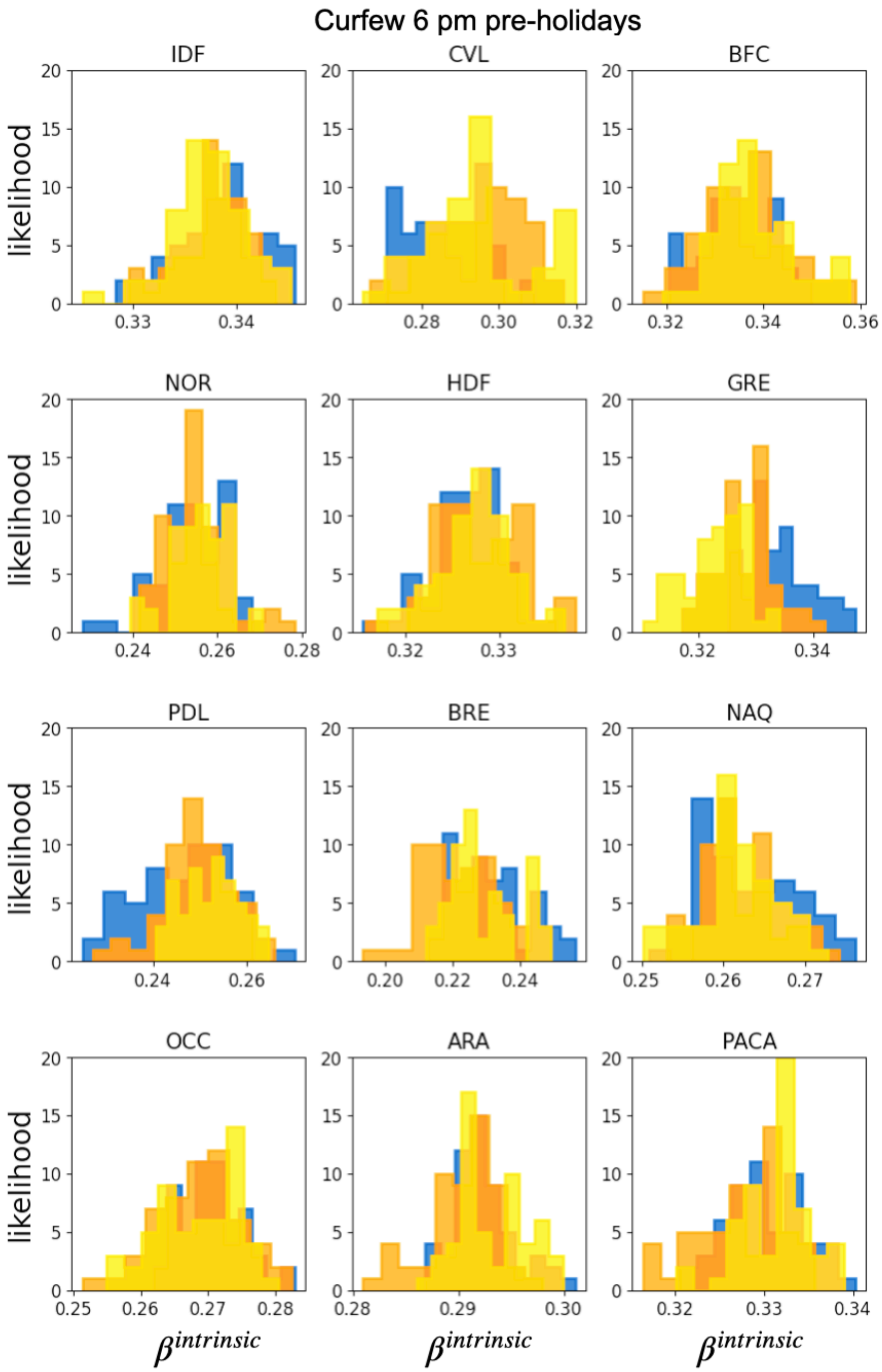

**Figure S6. Histograms of samples.** We reported the histograms of sampled values for the $\beta^{intrinsic}(curfew\ 6pm\ pre\ holidays)$ parameter in each region, obtained from three independent chains (yellow, orange and blue histogram), after discarding the burn-in period.

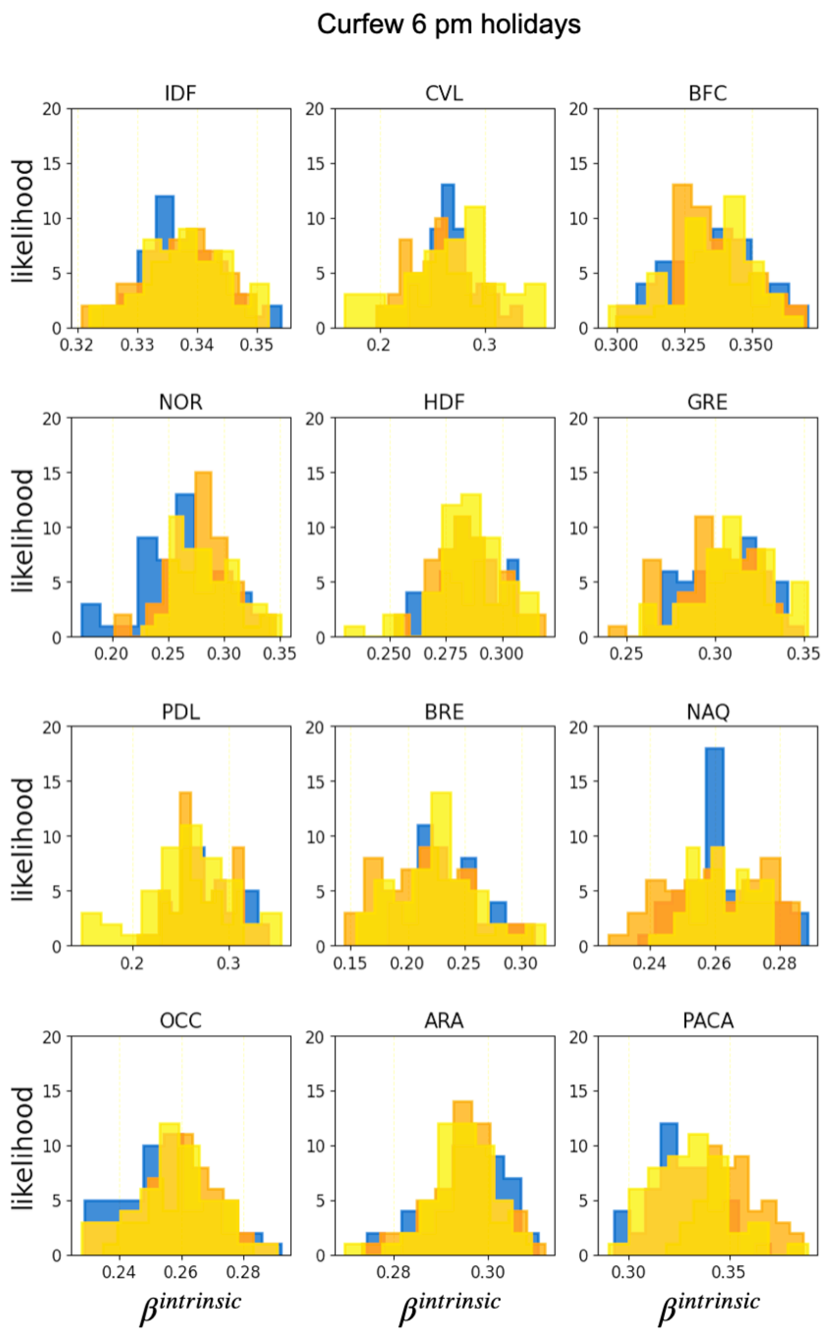

**Figure S7. Histograms of samples.** We reported the histograms of sampled values for the $\beta^{intrinsic}(curfew\ 6pm\ holidays)$ parameter in each region, obtained from three independent chains (yellow, orange and blue histogram), after discarding the burn-in period.

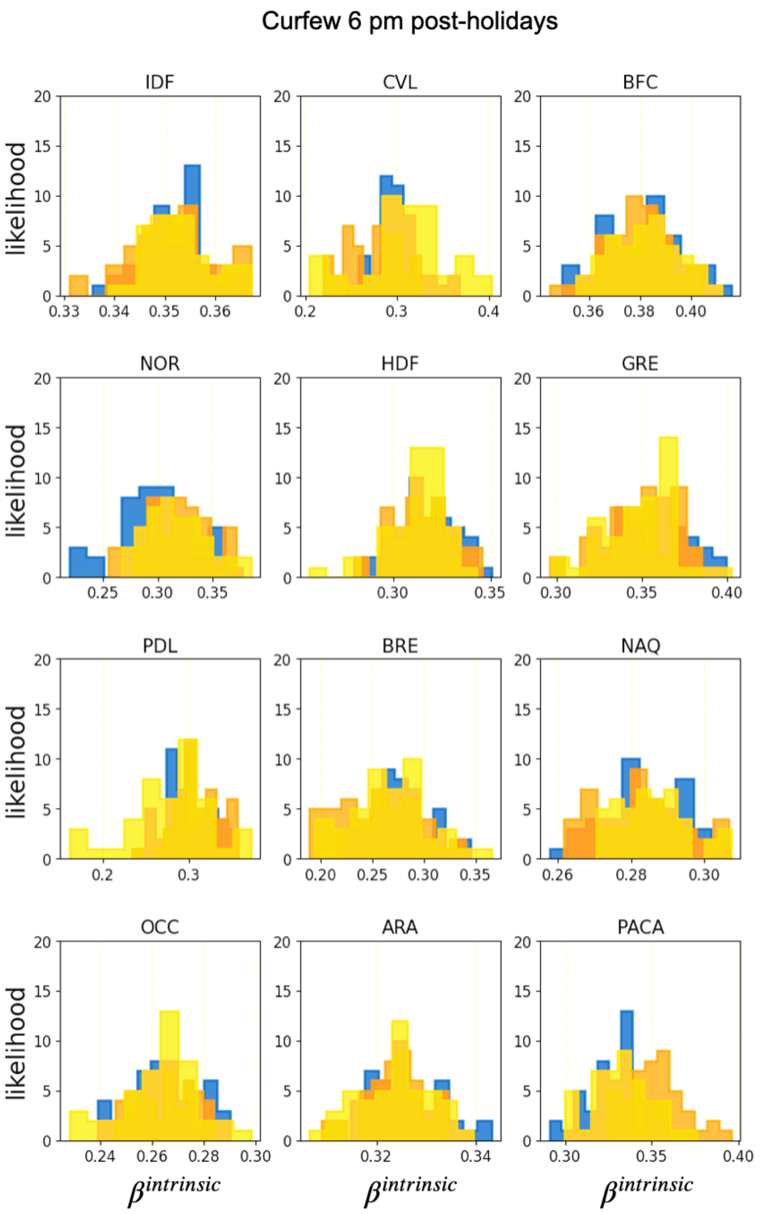

**Figure S8. Histograms of samples.** We reported the histograms of sampled values for the $\beta^{intrinsic}(curfew\ 6pm\ post\ holidays)$ parameter in each region, obtained from three independent chains (yellow, orange and blue histogram), after discarding the burn-in period.

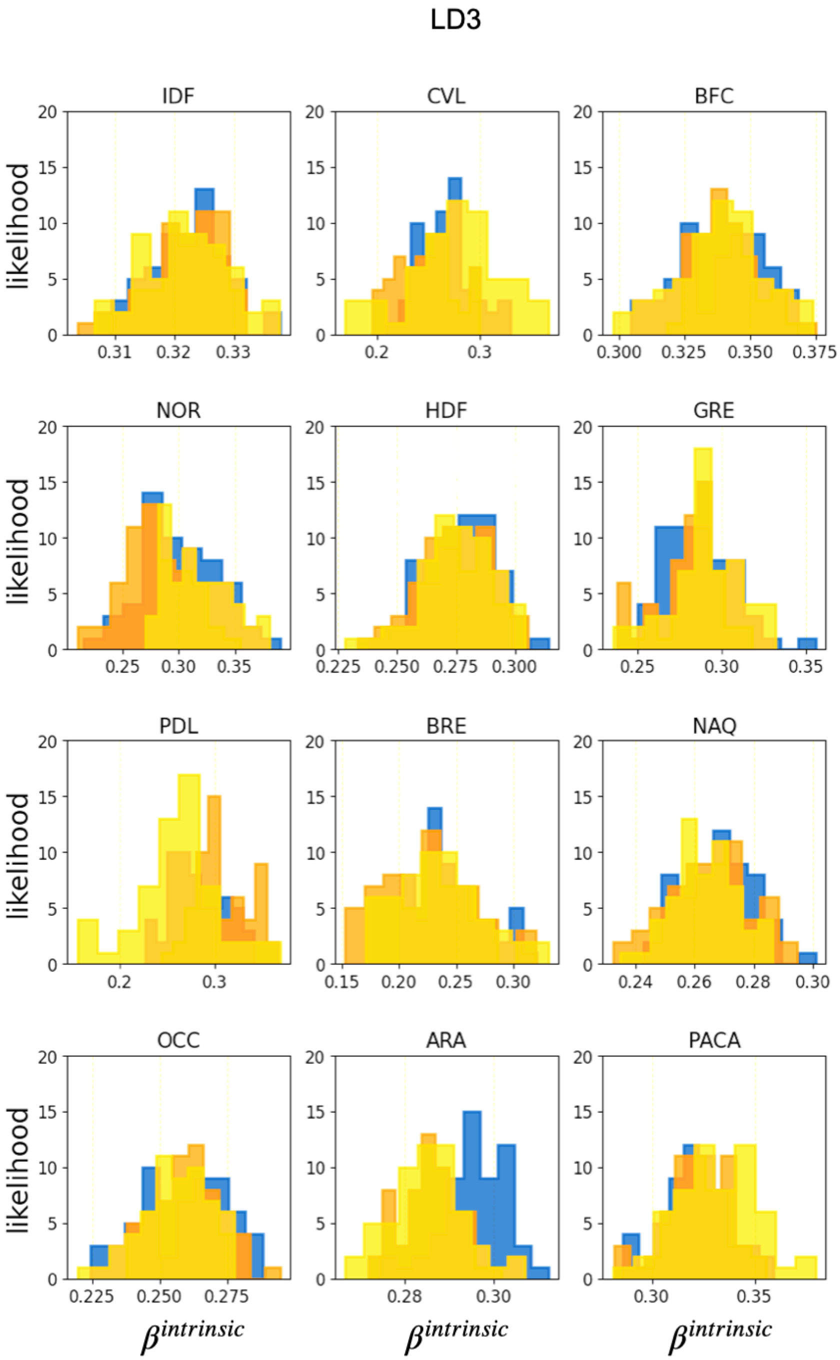

**Figure S9. Histograms of samples.** We reported the histograms of sampled values for the $\beta^{intrinsic}(curfew\ 6pm\ post\ holidays)$ parameter in each region, obtained from three independent chains (yellow, orange and blue histogram), after discarding the burn-in period.

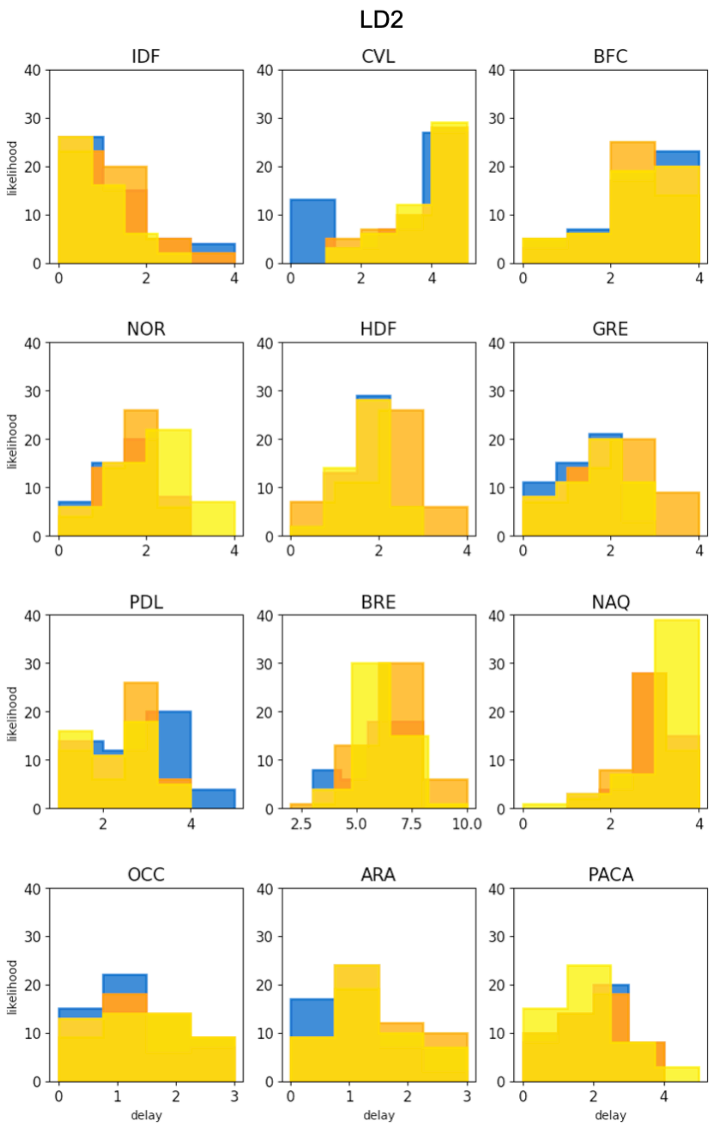

**Figure S10. Histograms of samples.** We reported the histograms of sampled values for the $delay(LD2)$ parameter in each region, obtained from three independent chains (yellow, orange and blue histogram), after discarding the burn-in period.

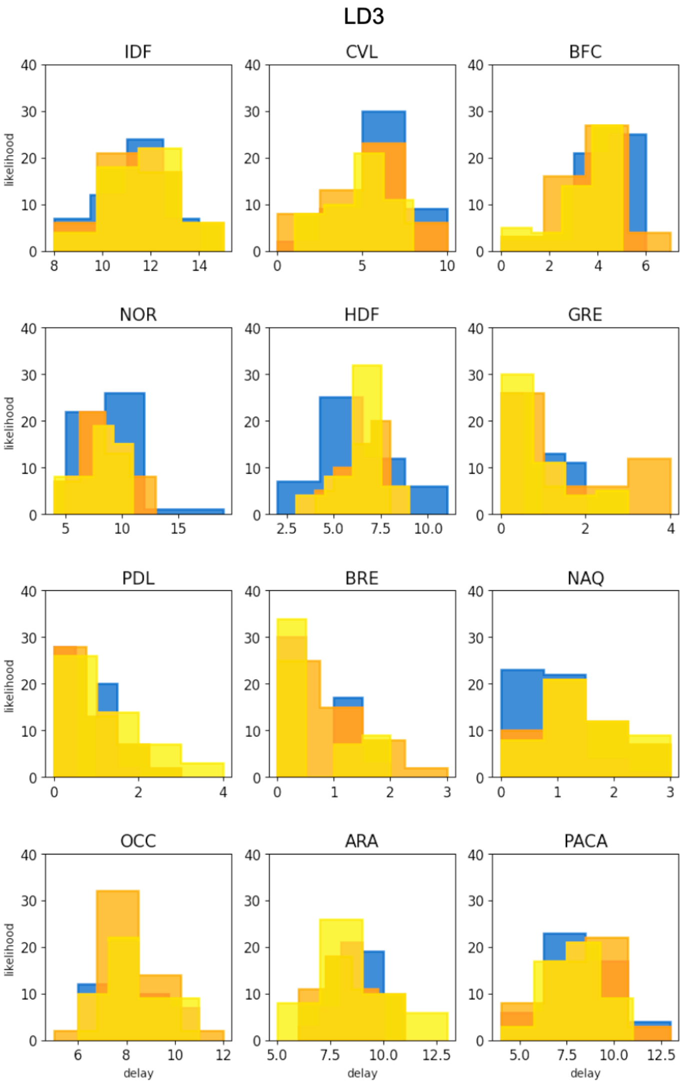

**Figure S11. Histograms of samples.** We reported the histograms of sampled values for the $delay(LD3)$ parameter in each region, obtained from three independent chains (yellow, orange and blue histogram), after discarding the burn-in period.

### 2.3. Model inputs

In **Figure S12** we show the model inputs, including the change in seasonality over time, the percentage of vaccine doses administrated over time, the mobility reduction, and the Alpha variant penetration over time. All these inputs were used in the model at the regional level.

Estimates from data from Ref.[14] on the impact of seasonal climatic conditions on transmissibility yield an average increase of 23% of transmissibility during winter. A stronger seasonal effect is observed in northern areas with respect to the south, with a maximum difference of 17 days in the winter peak observed across regions (**Figure S12a**). We fitted the estimates with a sinusoidal curve, one for each region, using a least-squares optimization function.

A large-scale vaccination campaign started in France on December 27, 2020, prioritizing the population at risk (elderly, vulnerable individuals and healthcare personnel). Data on the number of vaccine doses administered[15] included information at the regional level and by stage of vaccination (1st or 2nd dose). By June 12, 2021, around 30 million first injections had been distributed [16], corresponding to 45% of the total population and 20.8% of the total population was fully vaccinated with a second dose (**Figure S12b**). For comparison, we also tested scenarios following the vaccination pace adopted in the United Kingdom[17], where the fraction of population with a first dose reached 45% by April 15, 2021.

Different protocols were adopted over time for genome sequencing surveys to assess variant circulation. Flash#1 and Flash#2 surveys analyzed PCR-positive samples[18]. Flash#1 was conducted on Jan 8, identifying 3.3% of new cases due to Alpha. Flash#2 analyzed 10,261 samples from Jan 27, identifying 261 samples that were confirmed as Alpha variant (13.0%). Due to the need for more timely variant surveillance, a new protocol was introduced in week 6, 2021. It estimated the weekly frequency of detected viruses with specific mutations, including the N501Y mutation found in the Alpha variant, using second-line RT-PCR tests. Variant penetration was estimated by fitting the Flash surveys data[18] with a logistic function **(Figure S12d)**. To quantify the transmission advantage, we first estimated the daily effective reproductive numbers independently for each strain (wildtype or Alpha) at the national level starting on week 5, 2021. We then fitted the daily ratio $\frac{R_{Alpha}}{R_{wildtype}}$ with a zero-degree and a second-degree polynomial over time to allow possible variations of the transmission advantage over time. In agreement with prior estimates[18], we found that initially the SARS-CoV-2 Alpha variant was 1.58 times more transmissible than the wild type. The magnitude of the transmission advantage varied over time, decreasing from 1.58 in week 5, 2021 to 1.42 in week 22, 2021, similarly to what observed in United Kingdom[19]. These effects could be associated with vaccination, as vaccines may reduce outward transmission by reducing viral loads[13,19].

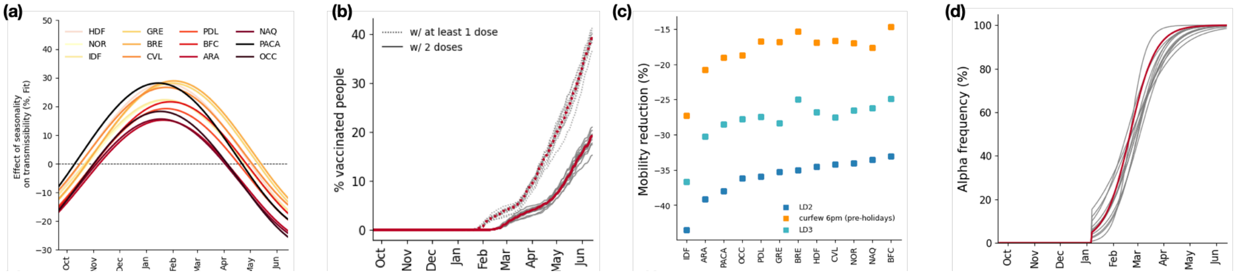

**Figure S12. Model inputs and indicators. (a)** Estimated effect of seasonality on the transmission over the study period. Regions are represented with solid lines of different colors: Île-de-France (IDF), Centre-Val de Loire (CVL), Bourgogne-Franche-Comté (BFC), Normandy (NOR), Hauts-de-France (HDF), Grand Est (GRE), Pays de la Loire (PDL), Brittany (BRE), Occitanie (OCC), Nouvelle Aquitaine (NAQ), Auvergne-Rhône-Alpes (ARA), Provence-Alpes-Cote d'Azur (PACA). **(b)** Percentage of vaccinated people with at least one dose (dotted lines) and with two doses (solid lines) according to data[15]. Grey lines represent the twelve French regions, red lines represent France. **(c)** Estimated change in mobility by region and by intervention based on Orange mobility data[20]. Dark blue squares represent the second lockdown period, light blue squares represent the third lockdown period and orange squares represent the curfew implemented at 6p.m. before the school holidays. **(d)** Percentage of Alpha variant over time. Grey lines represent the twelve French regions, red line represents France.

#### 2.4. Model validation

By including the processes of seroconversion and seroreversion following estimates of Ref.[21],we compared model projections of antibody positive people (AB+) with serological estimates[22,23](**Figures S13, S14, Methods** section). Modelling results are in good agreement with the serological estimates in the large majority of the regions and for the whole France.

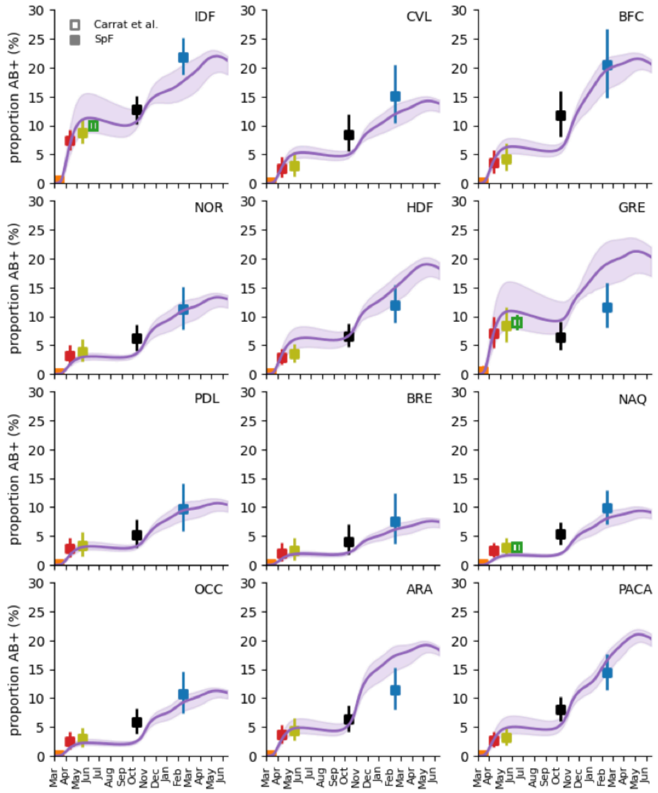

**Figure S13 Model predictions versus serological estimates, french regions**. For each region the panel shows the predicted percentage of antibody positive people (AB+) over time (purple curves and shaded areas for median and 95% probability range) and serological estimates of, Santé publique France (SpF)[22] (filled squares) and ref.[23] (void squares in IDF, GRE, NAQ). The square's colors refer to the different dates in which studies were conducted. Medians and 95% confidence intervals for model projections are obtained from n = 200 independent stochastic runs. Plots are reported for all 12 regions of mainland France: Île-de-France (IDF), Centre-Val de Loire (CVL), Bourgogne-Franche-Comté (BFC), Normandy (NOR), Hauts-de-France (HDF), Grand Est (GRE), Pays de la Loire (PDL), Brittany (BRE), Occitanie (OCC), Nouvelle Aquitaine (NAQ), Auvergne-Rhône-Alpes (ARA), Provence-Alpes-Cote d'Azur (PACA).

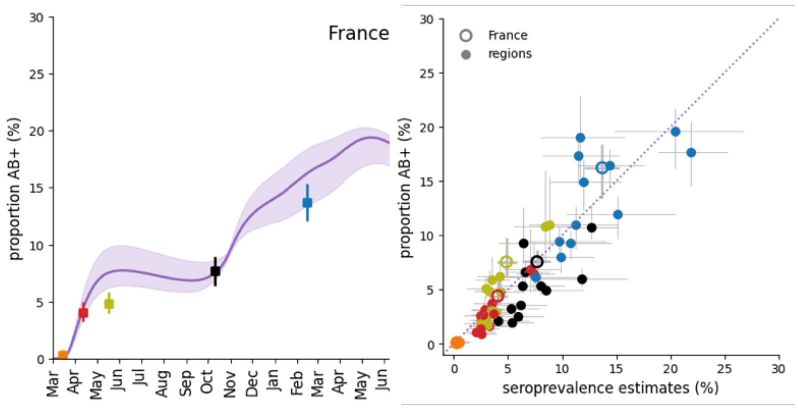

**Figure S14 Model predictions versus serological estimates, nationwide estimates**. (left) Model predicted percentage of antibody positive people (AB+) over time for France (purple curves and shaded areas for median and 95% probability ranges) and serological estimates of Santé publique France (SpF)[22], the square's colors refer to the different dates studies were conducted. (right) Model prediction of the percentage of antibody positive people versus the serological estimates per region (filled dots) and France (void dots) from Santé publique France (SpF)[22]. Error bars correspond to 95% probability ranges. The circles's colors refer to the different dates in which studies were conducted.

### 2.5. Spatial vs. Non-spatial model

We compared our metapopulation model with a non-spatial one, by fitting the transmission rates separately for each model, with and without spatial dependence. The model without spatial dependence considers that regions are not coupled by mobility. The resulting Deviance information criterion (DIC) shows that our model better describes the observed trajectories (**Table S4**, **Figure S15**), thus indicating that accounting for connections between regions is important to capture the epidemic dynamics.

**Table S4.** Deviance information criterion (DIC) values values for the two versions of the model.

| | **Metapopulation model** (i.e. model used in the study) | **Non-spatial model** |
|---|---|---|
| **DIC** | 36642.16 | 51536.97 |

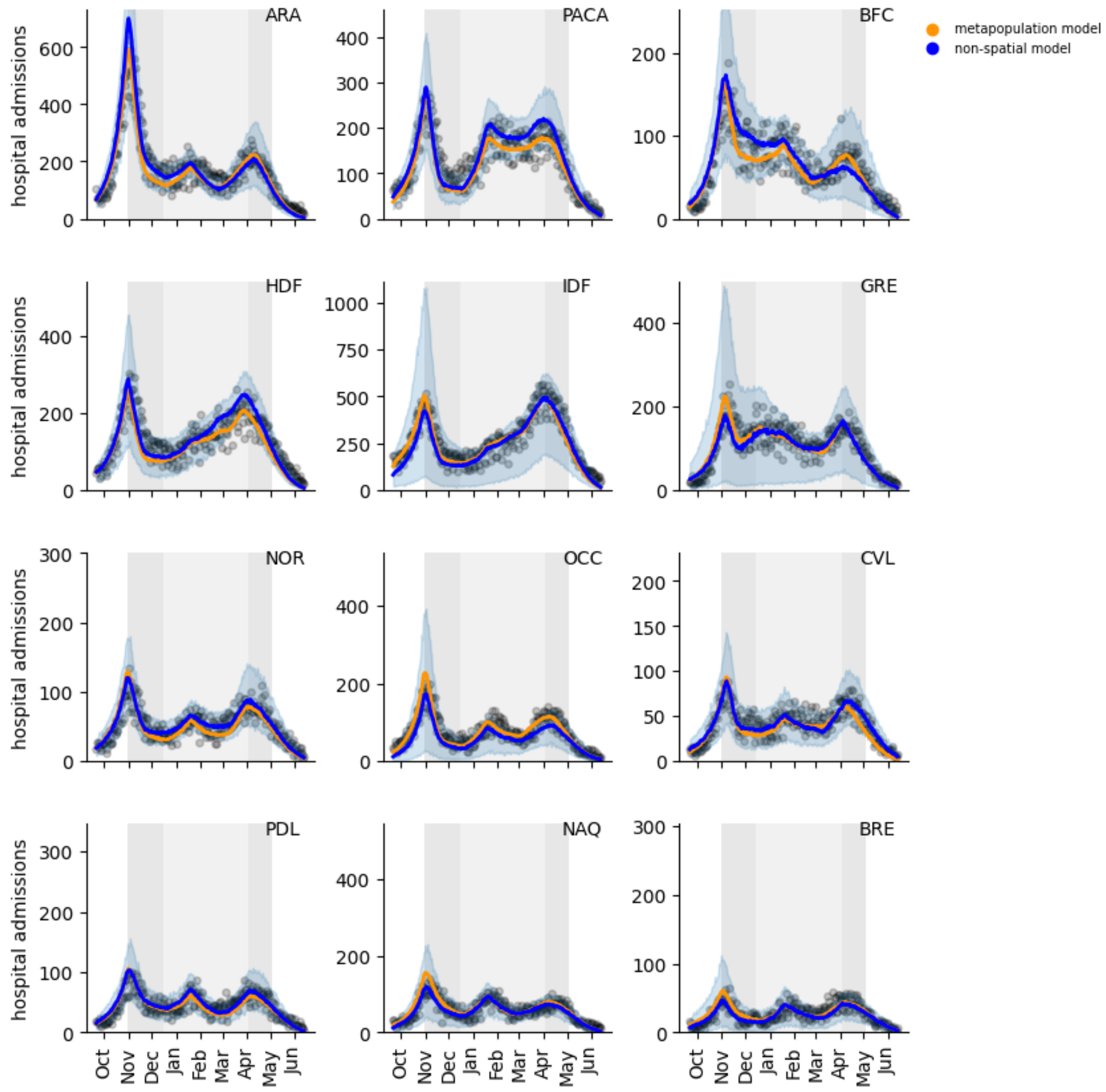

**Figure S15. COVID-19 pandemic trajectory in French regions: comparison between metapopulation model used in the study and the non-spatial model.** For each region, the panel shows the calibration of the model on data of daily hospital admissions with the metapopulation model (orange) and the non-spatial model (blue). Black dots indicate data, lines represent the median curve, shaded areas correspond to 95% probability ranges. Medians and 95% probability ranges for model projections are obtained from 200 independent stochastic runs. The abbreviations in the upper right corner of each plot stand for the name of the region. ARA : Auvergne-Rhône-Alpes, PACA : Provence-Alpes-Cote d'Azur, BFC : Bourgogne-Franche-Comté, HDF : Hauts-de-France, IDF: Île-de-France, GRE: Grand Est, NOR: Normandy, OCC : Occitanie, CVL: Centre-Val de Loire, PDL : Pays de la Loire, NAQ: Nouvelle Aquitaine, BRE: Brittany. Grey areas in the plots correspond to social distancing measures: lockdown during the second wave, lockdown during the third wave, and curfew in between.

We also computed the mean absolute error (MAE) for each run of each model. We obtained a lower MAE for the spatial model in 7 regions out of 11 (64%; for IDF the errors are compatible), **Figure S16**.

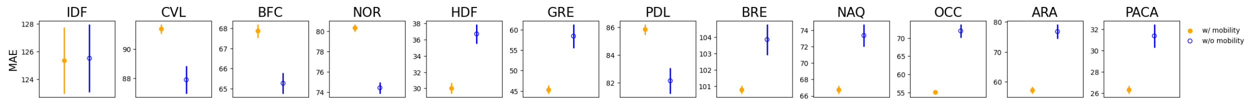

**Figure S16. MAE: comparison between metapopulation model used in the study and the non-spatial model.** For each region, the panel shows the mean absolute error. Circles represent the averaged MAE and its standard error estimated values for the 200 independent stochastic runs.

Filled circles represent the estimates with the metapopulation model; void circles represent estimates with the non-spatial one. Plots are reported for all 12 regions of mainland France : Île-de-France (IDF), Centre-Val de Loire (CVL), Bourgogne-Franche-Comté (BFC), Normandy (NOR), Hauts-de-France (HDF), Grand Est (GRE), Pays de la Loire (PDL), Brittany (BRE), Occitanie (OCC), Nouvelle Aquitaine (NAQ), Auvergne-Rhône-Alpes (ARA), Provence-Alpes-Cote d'Azur (PACA).

### 3. Counterfactual scenarios

**Table S5** and **Table S6** provided additional information on counterfactual scenarios. **Table S5** reports the values for the trigger (*T*) and release (*R*) thresholds of simulated lockdowns expressed in terms of daily hospital admissions per 100,000. **Table S6** reports the days spent under restrictions, for both the observed situation and the counterfactual scenarios discussed in the main text.

**Table S5.** Reference thresholds for trigger and release stop-and-go lockdowns. Values refer to daily hospital admissions per 100,000.

| Trigger threshold (T) | Release threshold (R) |
|---|---|
| 6.325 | 0.650 |

**Table S6.** Days spent under lockdowns.

| | Overall days | Effective days |
|---|---|---|
| observed | 227 (47 in LD2, 149 under curfew, 31 in LD3) | 157.291 |
| T-70%R-35% | 196 (over 3 lockdowns) | 156.51 |
| T-35%R-70% | 189 (over 2 lockdowns) | 154.84 |
| T,R | 150 (over 3 lockdowns) | 122.28 |

### 4. Additional results

We present in this section some additional results of our analyses not shown in the main text.

#### 4.1. Estimated impact of implemented NPIs on the reproductive number

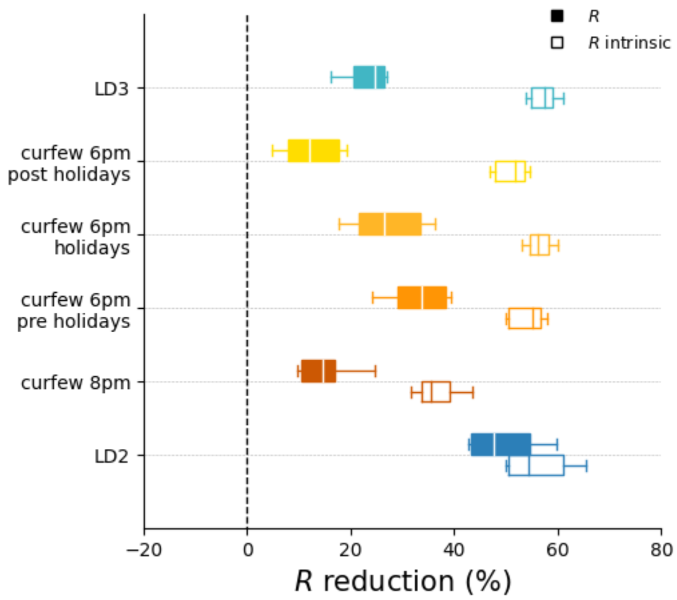

**Figure S17. Estimated impact of implemented NPIs.** Reduction in the estimated effective reproductive numbers R associated to the implemented social distancing interventions compared with the values estimated before the second lockdown. Box plots represent the median (line in the middle of the box), interquartile range (box limits) and 2.5th and 97.5th percentiles (whiskers) of the estimated values for the 12 French regions. Filled boxplots represent reductions estimated by the fit accounting for all time-varying processes; void boxplots represent reductions estimated in the absence of the Alpha variant and seasonality effects.

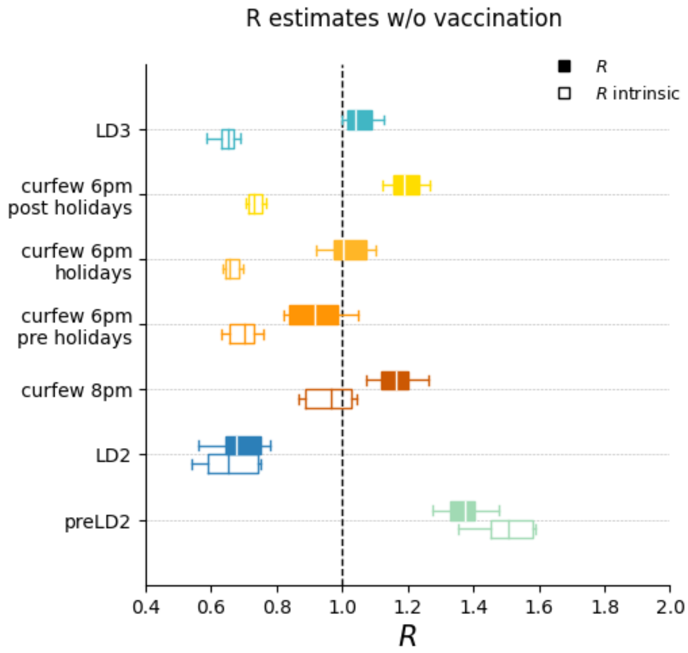

**Figure S18. Estimated impact of implemented NPIs, in absence of vaccination.** Estimates of the effective reproductive numbers for the implemented social distancing interventions in absence of vaccination. Box plots represent the median (line in the middle of the box), interquartile range (box limits) and 2.5th and 97.5th percentiles (whiskers) of the estimated values for the 12 French regions. Filled boxplots represent reductions estimated by the fit accounting for all time-varying processes; void boxplots represent reductions estimated in the absence of the Alpha variant and seasonality effects.

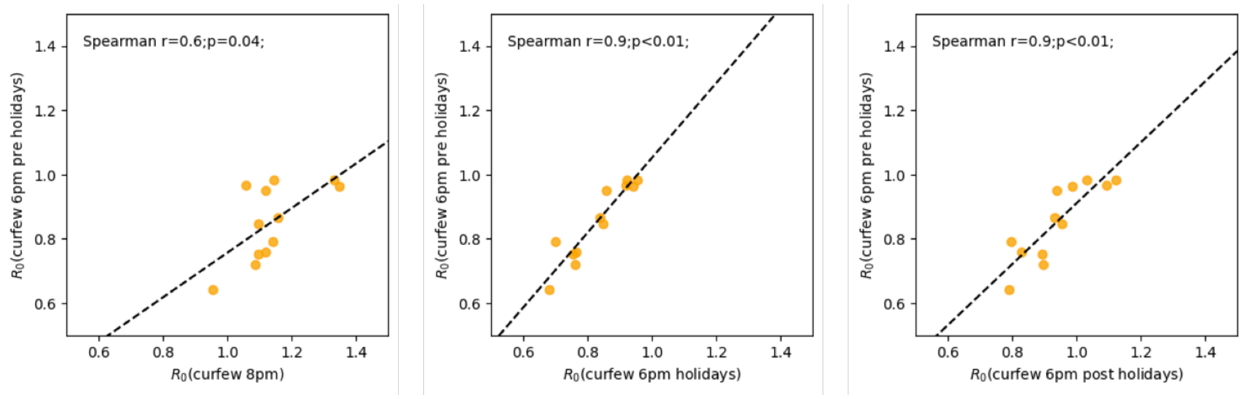

**Figure S19. Correlations between basic reproductive numbers during different interventions.** The panel shows the correlation of the basic reproductive number $R_0$ during the period with curfew at 6p.m. before holidays and curfew at 8 p.m., curfew at 6 p.m. during holidays, curfew at 6 p.m. after holidays. Each dot represents a French region and correlation is done across regions.

#### 4.2. Impact of the Alpha variant and of the vaccination rhythm on the hospitalizations

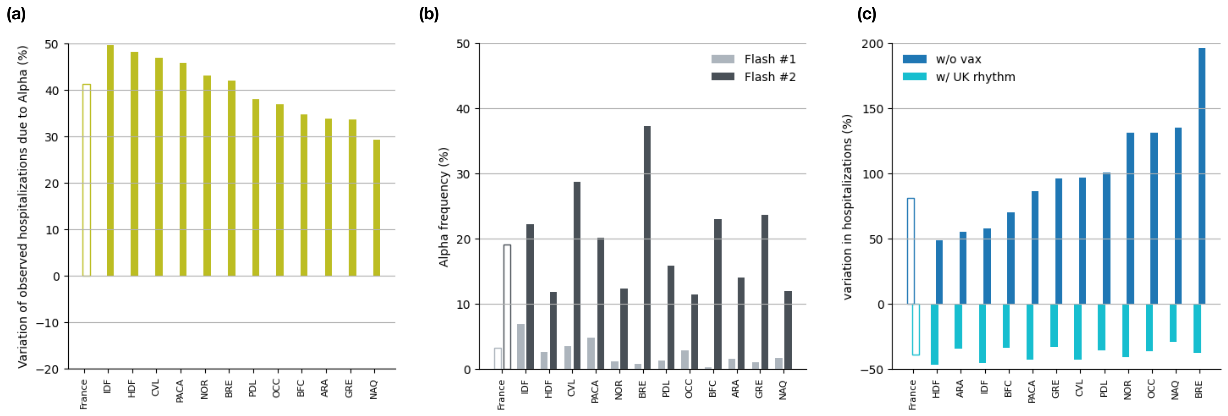

**Figure S20. Impact of the Alpha variant and of the vaccination rhythm on hospitalizations. (a)** Variation in the number of hospitalizations (period September 2020 - June 2021) due to the Alpha variant with respect to observations. The green bars represent a scenario without the Alpha variant. **(b)** Frequency of Alpha variant (%), by region according to Flash surveys . The light grey bars represent Flash #1, the dark grey bars represent Flash #2. **(c)** Variation in the number of hospitalizations (period September 2020 - June 2021) due to the vaccination rhythm compared to observations. The dark blue bars represent a scenario without vaccines, the light blue bars represent a context with the vaccination pace observed in the United Kingdom[17]. In the three panels, the empty bars represent values for France, the filled bars represent the regional values.

#### 4.3. Impact of different nationwide interventions

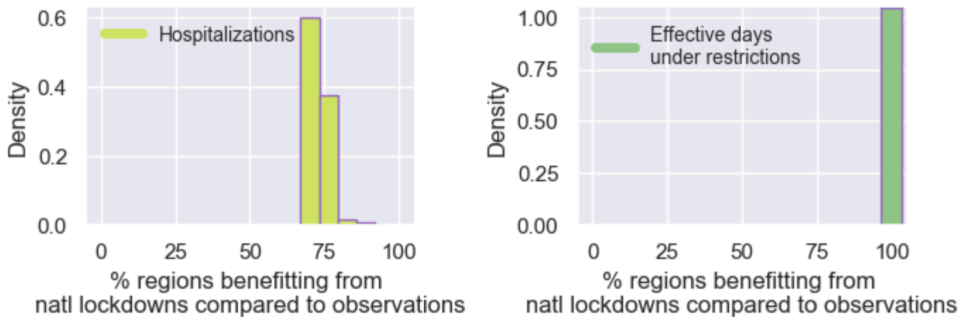

**Figure S21. Regions benefitting from nationwide lockdowns compared to observations.** Probability distribution of the percentage of regions benefitting from the lockdowns compared to observations, in the phase space where both effective days and hospitalizations are reduced. From left to right: hospitalizations, effective days under restrictions.

### 5. Sensitivity analysis

Here we present the results of our sensitivity analysis on some assumptions considered in the model used in the main paper. We used the value T,R of trigger and release described in the main text, as an illustrative example.

#### 5.1. Impact of relaxation after exiting lockdowns

We assume that population behavior did not change immediately with policies: the population continued to adhere to public health measures being progressively lifted, such as physical distancing, during the reopening phase. We assume two weeks relaxation in the main text. Other studies[24] found that transmission rate during the reopening phases following the lifting of lockdowns remained similar to the one observed during lockdowns. Here we explore one week relaxation or no relaxation (**Figure S22**), and we find that the progressive reopening helps further dampening the waves over time.

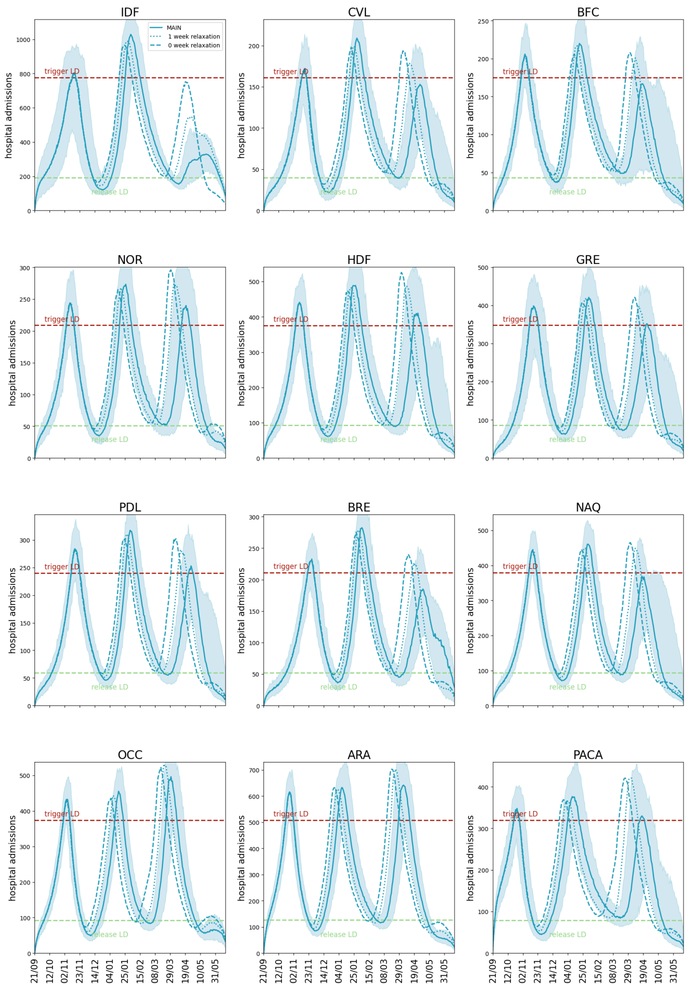

**Figure S22. Impact of relaxation after exiting lockdowns on daily hospital admissions.** Solid curves refer to the median trajectory, obtained under the condition of 2 weeks relaxation. Dotted and dashed curves show the same for 1 week relaxation o no relaxation, respectively. The shaded area around the curves corresponds to the 95% probability range obtained from n=200 stochastic simulations. The abbreviations in the upper right corner of each plot stand for the name of the region. IDF: Île-de-France, CVL: Centre-Val de Loire, BFC : Bourgogne-Franche-Comté, NOR: Normandy, HDF: Hauts-de-France, GRE: Grand Est, PDL : Pays de la Loire, BRE: Brittany, NAQ: Nouvelle Aquitaine, OCC : Occitanie, ARA: Auvergne-Rhône-Alpes, PACA: Provence-Alpes-Côte d'Azur. Dashed horizontal red lines refer to the trigger threshold relative to the second lockdown. It is estimated based on the 7-days rolling average value of the hospital admissions per capita in the Auvergne-Rhone-Alpes region, on October 30, 2020, rescaled by the regional population, the green ones to the ones observed on December 15, 2020, calculated as the average value between regions and rescaled to the region's population.

### 5.2. Impact of transmissibility and mobility conditions during lockdowns

In the main text, we simulated the stringency and mobility reductions experienced in the French second lockdown (LD2) for the initial lockdown in the stop-and-go series, and the stringency and mobility reductions experienced in the French third lockdown (LD3) for the subsequent simulated lockdowns. This was done to align with the applied policies aiming towards a larger freedom over time. Here we show the results using only the lockdown as the second phase (**Figure S23**). Assuming that all simulated lockdowns in the stop-and-go series have the same stringency and mobility reduction as the French second lockdown does not bring substantial changes to our results. This is due to the similarity of estimated effectiveness of LD2 and LD3.

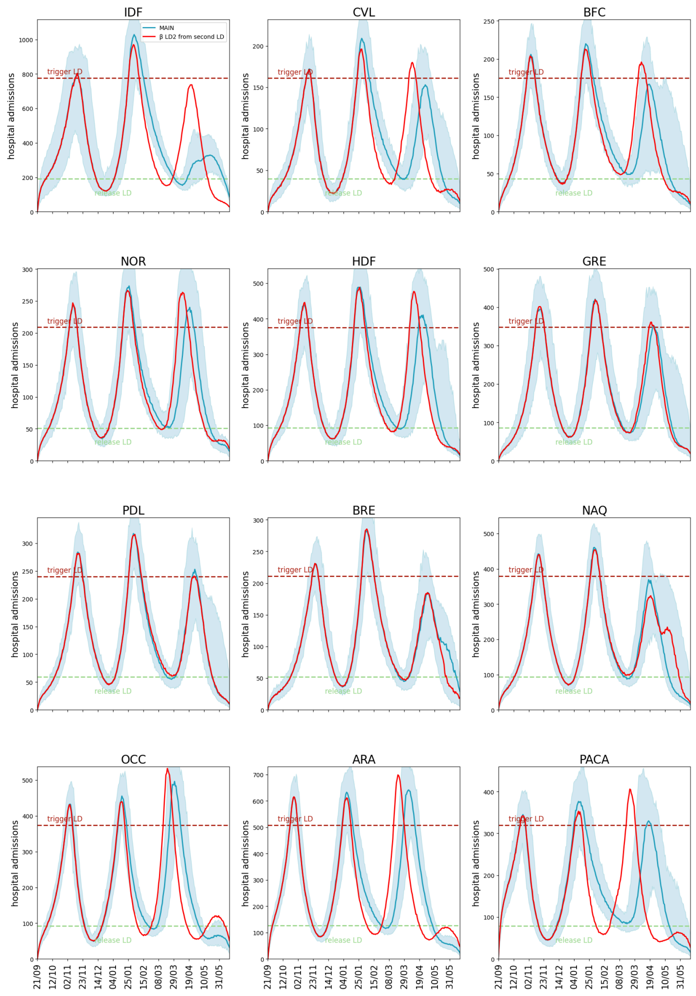

**Figure S23. Impact of transmissibility and mobility conditions during lockdowns on daily hospital admissions.** Blue solid curves refer to the median trajectory, obtained under the main scenario conditions. Red solid curves show the same assuming the conditions of the second lockdown for all the epidemic waves. The shaded area around the curves corresponds to the 95% probability range obtained from n=200 stochastic simulations. The abbreviations in the upper right corner of each plot stand for the name of the region. IDF: Île-de-France, CVL: Centre-Val de Loire, BFC : Bourgogne-Franche-Comté, NOR: Normandy, HDF: Hauts-de-France, GRE: Grand Est, PDL : Pays de la Loire, BRE: Brittany, NAQ: Nouvelle Aquitaine, OCC : Occitanie, ARA: Auvergne-Rhône-Alpes, PACA: Provence-Alpes-Côte d'Azur. Dashed horizontal lines refer to the hospitalization per capita. Dashed horizontal red lines refer to the trigger threshold relative to the second lockdown. It is estimated based on the 7-days rolling average value of the hospital admissions per capita in the Auvergne-Rhone-Alpes region, on October 30, 2020, rescaled by the regional population, the green ones to the ones observed on December 15, 2020, calculated as the average value between regions and rescaled it to the region's population.

### 5.3. Impact of weekly rolling average of data

We show that fitting the model to the weekly rolling average of the hospital admission data does not alter the estimates of the epidemiological impact of the NPIs (**Figure S24**).

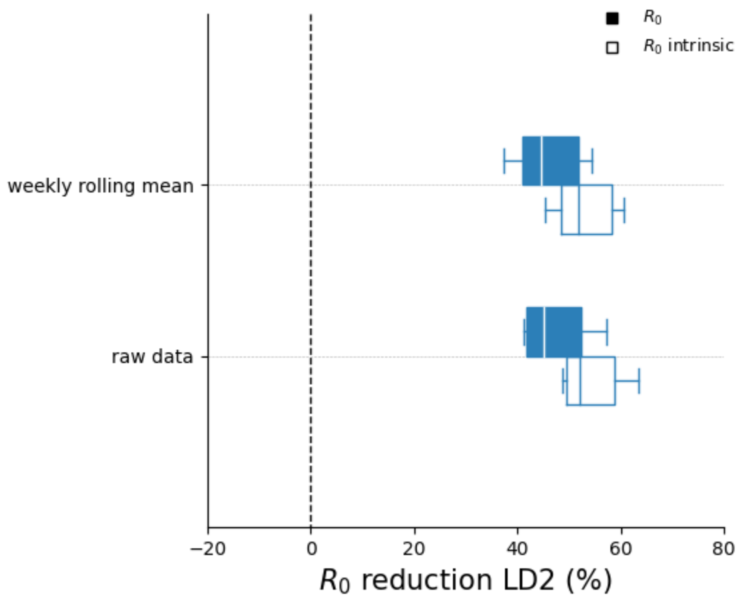

**Figure S24. Impact of hospitalization data on NPI effectiveness.** Reduction in the estimated regional basic reproductive numbers $R_0$ associated to the second lockdown (LD2) compared with the values estimated before the second lockdown using raw data and a weekly average of the data. Box plots represent the median (line in the middle of the box), interquartile range (box limits) and 2.5th and 97.5th percentiles (whiskers) of the estimated values for the 12 French regions. Filled boxplots represent reductions estimated by the fit accounting for all time-varying processes ($R_0$); void boxplots represent the same reductions discounting the seasonal and Alpha effects ($R_0^{intrinsic}$).
